# A blueprint for academic labs to produce SARS-CoV-2 RT-qPCR test kits

**DOI:** 10.1101/2020.07.29.20163949

**Authors:** Samantha J. Mascuch, Sara Fakhretaha-Aval, Jessica C. Bowman, Minh Thu H. Ma, Gwendell Thomas, Bettina Bommarius, Chieri Ito, Liangjun Zhao, Gary P. Newnam, Kavita R. Matange, Hem R. Thapa, Brett Barlow, Rebecca K. Donegan, Nguyet A. Nguyen, Emily G. Saccuzzo, Chiamaka T. Obianyor, Suneesh C. Karunakaran, Pamela Pollet, Brooke Rothschild-Mancinelli, Santi Mestre-Fos, Rebecca Guth-Metzler, Anton V. Bryksin, Anton S. Petrov, Mallory Hazell, Carolyn B. Ibberson, Petar I. Penev, Robert G. Mannino, Wilbur A. Lam, Andrés J. Garcia, Julia M. Kubanek, Vinayak Agarwal, Nicholas V. Hud, Jennifer B. Glass, Loren Dean Williams, Raquel L. Lieberman

## Abstract

Widespread testing for the presence of the novel coronavirus SARS-CoV-2 in individuals remains vital for controlling the COVID-19 pandemic prior to the advent of an effective treatment. Challenges in testing can be traced to an initial shortage of supplies, expertise and/or instrumentation necessary to detect the virus by quantitative reverse transcription polymerase chain reaction (RT-qPCR), the most robust, sensitive, and specific assay currently available. Here we show that academic biochemistry and molecular biology laboratories equipped with appropriate expertise and infrastructure can replicate commercially available SARS-CoV-2 RT-qPCR test kits and backfill pipeline shortages. The Georgia Tech COVID-19 Test Kit Support Group, composed of faculty, staff, and trainees across the biotechnology quad at Georgia Institute of Technology, synthesized multiplexed primers and probes and formulated a master mix composed of enzymes and proteins produced in-house. Our in-house kit compares favorably to a commercial product used for diagnostic testing. We also developed an environmental testing protocol to readily monitor surfaces across various campus laboratories for the presence of SARS-CoV-2. Our blueprint should be readily reproducible by research teams at other institutions, and our protocols may be modified and adapted to enable SARS-CoV-2 detection in more resource-limited settings.

## Introduction

The global COVID-19 pandemic caused by severe acute respiratory syndrome coronavirus 2 (SARS-CoV-2) substantially disrupted activities in the public and private sectors (1-3). Widespread and frequent testing, in conjunction with contact tracing and behavioral change, has been demonstrated by some countries to be effective in monitoring and managing the outbreak. These strategies will continue to be instrumental in containing the virus until a vaccine or other effective treatment is universally available (4). While comprehensive testing programs have been successfully implemented in many countries, testing efforts in the US were hampered by a lack of access and an uncoordinated approach to early testing.

The Georgia Tech COVID-19 Test Kit Support Group was conceived to leverage in-house Georgia Tech facilities, expertise, and personnel to assist State of Georgia Clinical Laboratory Improvement Amendments (CLIA) labs with materials needed for clinical SARS-CoV-2 detection. Similar efforts are underway at several other universities (5,6), with some going so far as to establish “pop-up” testing labs (7). To our knowledge, ours is the first effort to use all in-house materials and equipment, offering an open-access community resource for other settings with similar capabilities.

Much of the testing shortfall, especially early in the pandemic, can be traced to a shortage of reagents, plasticware, expertise, or instrumentation necessary to perform quantitative reverse transcription polymerase chain reaction (RT-qPCR (8)). In RT-qPCR, RNA is converted to cDNA which is then amplified via PCR until a detection threshold is reached. The TaqMan RT-qPCR method is widely considered the “gold standard” for SARS-CoV-2 testing due to its robustness, high sensitivity, linearity, and specificity (9). In a TaqMan RT-qPCR, the 5’ −3’ exonuclease activity of a thermostable DNA polymerase cleaves a TaqMan oligonucleotide probe hybridized to the PCR amplicon. One terminus of the TaqMan probe is linked to a fluorophore and the other terminus is linked to a quencher. Success in reverse transcription and PCR is detected as an increase in fluorescence upon probe cleavage during successive rounds of PCR, producing a sensitive and quantitative fluorescence signal that may be monitored in real time.

A complete TaqMan RT-qPCR test kit includes (i) solution(s) of matched DNA probe(s) and primers specific to the gene target(s) of interest and (ii) an enzyme master mix. These solutions are mixed with a sample suspected to contain the RNA (e.g., SARS-CoV-2 RNA), run through a thermal cycling protocol in an RT-qPCR instrument, and monitored for an increase in fluorescence indicative of the presence of the target RNA. Solutions of matched probe and primer can be designed to detect one (singleplex) or multiple (multiplex) targets in a single reaction. Detection of a target usually needs to be differentiable from other targets in a multiplex reaction. In this case, a distinct fluorophore, with non-overlapping emission wavelength, is used for each target. Commercial enzyme master mixes are sometimes branded for use with multiplex or singleplex primers and probes.

The original CDC SARS-CoV-2 assay (10) was a singleplex assay that required four distinct reactions for each sample suspected to contain the SARS-CoV-2 RNA: one for each of two sets of primers/probes (N1 and N2) targeting different regions of the N gene that encodes the SARS-CoV-2 nucleocapsid protein, one for a third set of primers/probe (N3) that detects all clade 2 and 3 viruses of the *Betacoronavirus* subgenus *Sarbecovirus*, and one for the primers/probe targeting human RNase P (RP). The latter is a control reaction for monitoring performance of the sample collection and RNA extraction. The CDC N3 primers/probe were later eliminated due template contamination and because they are unnecessary for specific detection of SARS-CoV-2 (10,11), leaving three distinct reactions per sample. All probes in the CDC singleplex assay bear the common FAM fluorophore. Many companies subsequently developed FDA-approved multiplex SARS-CoV-2 primer/probe sets with a variety of fluorophores, enabling detection of all targets in a single reaction. Compared to singleplex, use of multiplex primer/probe sets substantially reduces the amount of enzyme mix and plasticware needed to process one patient sample but requires RT-qPCR instrumentation capable of monitoring the specific combination of fluorophores used.

The TaqPath 1-Step RT-qPCR Master Mix (Thermo Fisher Scientific) was the first enzyme master mix to be recommended by the CDC and approved by the FDA for detection of SARS-CoV-2 (10). TaqPath is a “unique proprietary formulation” containing a thermostable MMLV reverse transcriptase, a fast and thermostable DNA polymerase, an RNase inhibitor, a heat labile uracil-N glycosylase (UNG), dNTPs including dUTP, ROX™ passive reference dye, and a buffer containing stabilizers and other additives. The DNA polymerase in TaqPath is likely to be a mutant of *Taq* polymerase incorporating some type of hot-start technology (12) to help suppress nonspecific amplification and primer dimers. UNG can remove carry-over contamination by specifically degrading products of prior PCRs that incorporate dUTP.

Three other enzyme mixes, two made by Quantabio (qScript XLT One-Step RT-qPCR ToughMix (2X) and UltraPlex 1-Step ToughMix (4X)), and one made by Promega (GoTaq® Probe 1-Step RT-qPCR System) were added to the CDC list of master mix options shortly after TaqPath. The Quantabio mixes are provided as single components at 2X or 4X concentration, each containing a reverse transcriptase, antibody-based hot-start *Taq* DNA polymerase, RNase inhibitor protein and the standard set of dNTPs. The Promega mix is provided as multiple components including a 2X mix containing an antibody-based hot-start *Taq* and dNTPs (including dUTP), and a 50X mix containing reverse transcriptase and recombinant RNase inhibitor.

Here we describe our in-house RT-qPCR assay for detection of SARS-CoV-2 (**Fig. 1**). First, we discuss preparation of singleplex and multiplex primers and probes with CDC sequences that can be used with commercial enzyme master mixes. Second, we present the production of reverse transcriptase (RT), *Taq* DNA polymerase, and ribonuclease inhibitor (RI) proteins, and the formulation of a working 1-step enzyme Georgia Tech master mix (GT-Master Mix) for use with our primers and probes. We compare the performance of our full in-house kit to a commercial kit. Finally, we describe implementation of environmental testing for SARS-CoV-2 across various campus laboratories.

**Figure 1.**
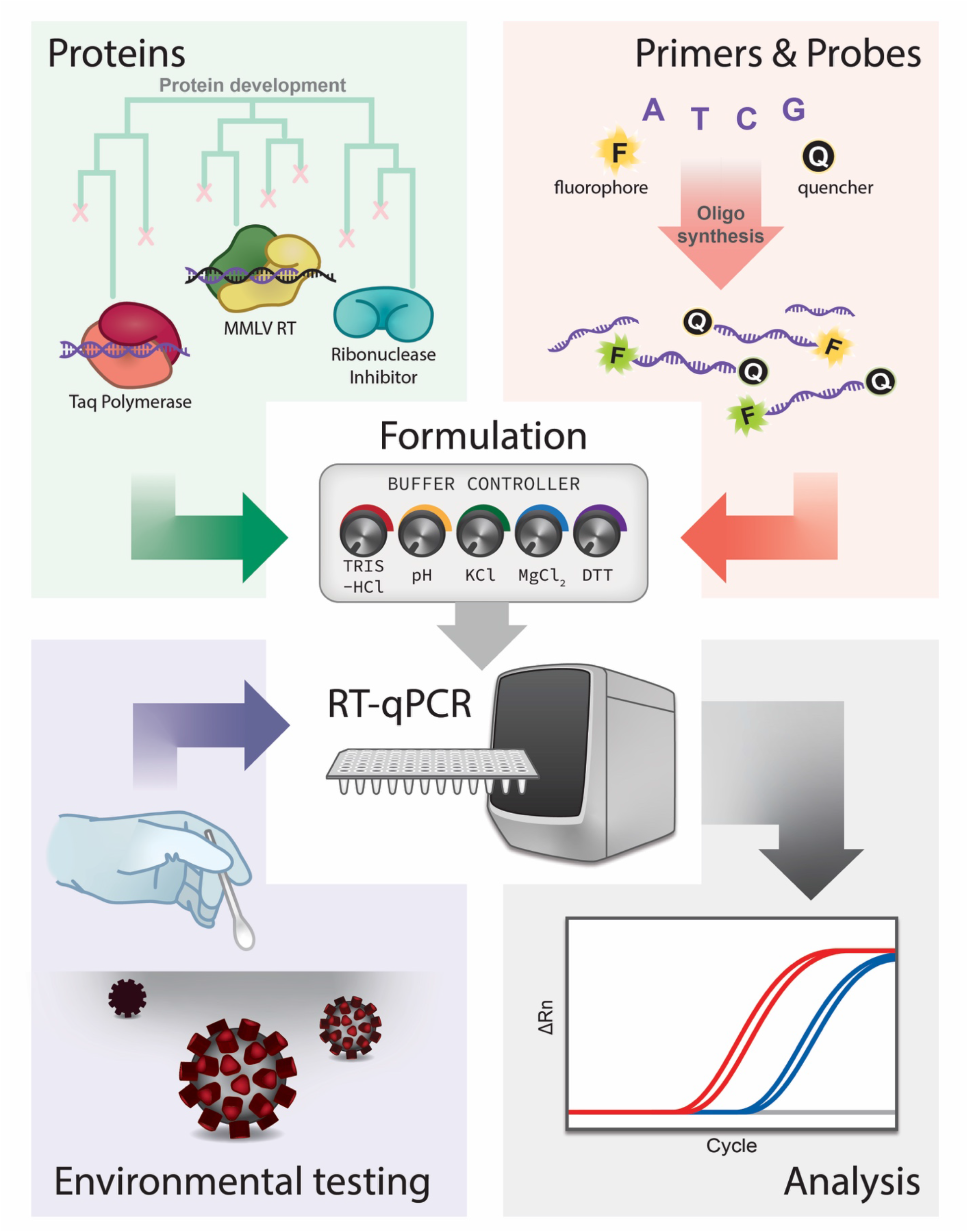
Project components and workflow.

## Results

### Primers and probes

We focused on producing N1 and N2 primer and probe system published by CDC in March of 2020 (**Table 1**) because (i) these sequences had been extensively verified in the literature, (ii) our own bioinformatics analysis showed them to be highly specific to SARS-CoV-2 and localized to regions of the genome with low mutation rates (not shown), and (iii) they had received FDA Emergency Use Authorization (EUA). First, we synthesized and assayed the same singleplex primers and probes specified by the CDC. We then converted the CDC singleplex probes and primers to a multiplex system, in which N1 and N2 are detected via a common channel, and RNase P is detected in a separate channel. Specifically, the CDC FAM-RP-BHQ1 probe was converted to HEX-RP-BHQ1 to allow its simultaneous detection alongside FAM-N1-BHQ1 and FAM-N2-BHQ1, and the three probe/primer sets were combined in a single solution. The HEX fluorophore (maximum λ_em_=556nm) is compatible with standard fluorophore channels of commercial RT-qPCR instruments and distinguishable from the FAM emission maximum at 518 nm. In addition, HEX has the second highest quantum yield (0.7) after FAM (0.9) and is commercially available. During development of our multiplex probe set, the OPTI SARS-CoV-2 RT-PCR multiplex test kit (13) which uses the same HEX-RP-BHQ1/FAM-N1-BHQ1/FAM-N2-BHQ1 probe configuration as our GT kit, gained EUA from the US FDA (May 2020).

**Table 1.**
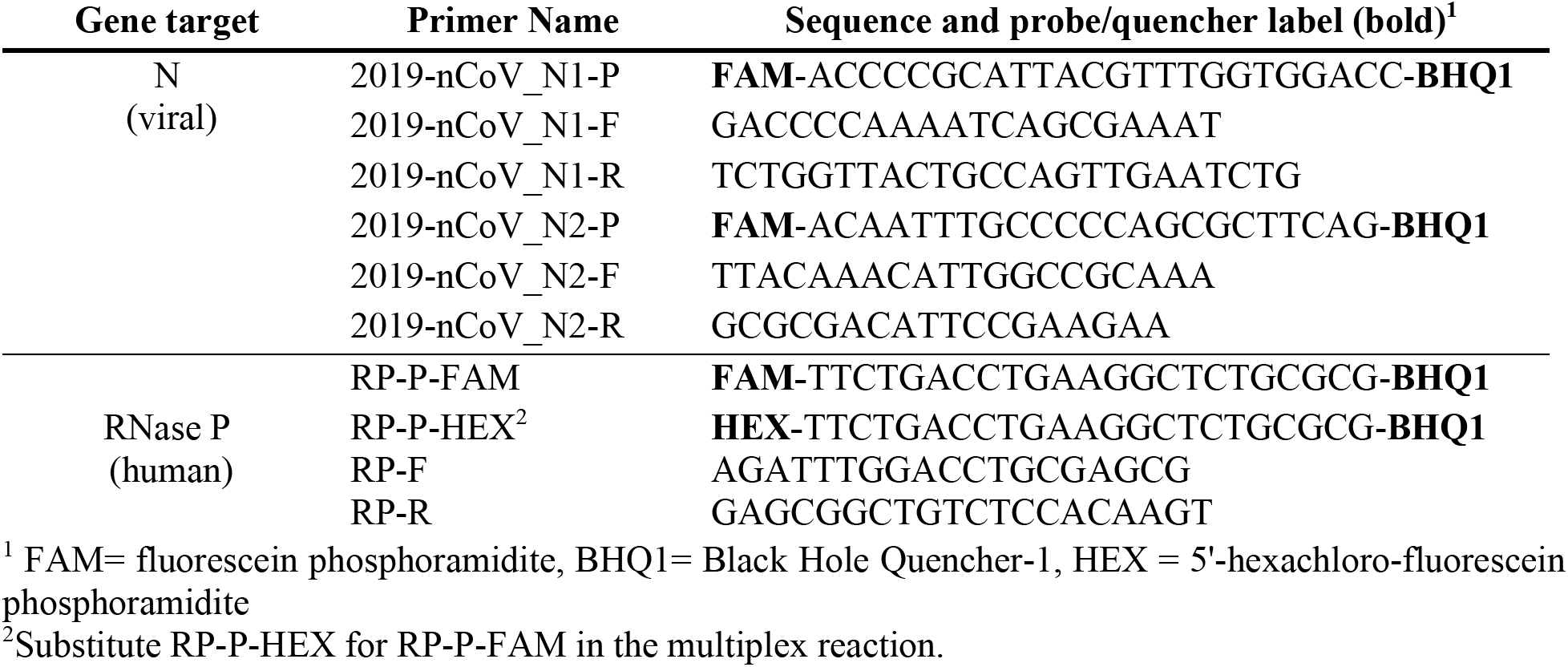
Sequences of CDC primers and probes. FAM was substituted for HEX in RP probe.

The GT Parker H. Petit Institute for Bioengineering and Bioscience’s Molecular Evolution Core Facility dedicated its ASM-2000 high throughput DNA/RNA synthesizer to primer and probe syntheses, which enabled tens of thousands of reactions worth of primers and probes to be produced in-house with a 6-8 h turn-around. Initially, HPLC was used to purify probes, but cross-contamination was detected from IDT positive control plasmid that was handled in the same laboratory (see details in environmental testing below). Learning from contamination issues faced by CDC (11), and to further avoid potential contamination across the multi-tasking academic labs, a cartridge method was subsequently used to polish the FAM probes in a separate academic building, and HPLC was only used to analyze purity of an aliquot of the material (Supporting Information (**Supp**.) **Fig. S1A-D**). Primer purity was confirmed by gel electrophoresis after ^32^P end-labeling (**Supp. Fig. S1E**). For the HEX probe, we used unpurified, but carefully synthesized HEX probe based on the literature precedent that it should achieve similar efficiency as the high purity probe (14). HEX probe purity ranged from 35-60% over different synthetic batches (not shown). The HEX probe exhibited temperature-dependent enhancement of fluorescence intensity, as expected (**Supp. Fig. S1F**).

GT-made primers and probes generated robust and reproducible RT-qPCR signals with their respective targets. Prior to use with GT-Master Mix, primers and probes were individually validated by RT-qPCR to ensure acceptable performance in detecting N1, N2, and/or RP targets in commercial master mix (**Supp. Fig. S2A-C**) and no contamination. Performance of FAM-labeled N1 and N2 is similar to commercial primers and probes purchased from IDT (not shown). The GT multiplex primers and probes mix was evaluated using several commercially available enzyme master mixes. The performance of multiplexed primers and probes did not differ among the enzyme master mixes tested (**Fig. 2**), and did not differ from the singleplex system (**Supp. Fig. S2C**). Thus, multiplexing did not impair enzyme function or deplete potentially limiting reagents, like dNTPs, from the master mix.

**Figure 2.**
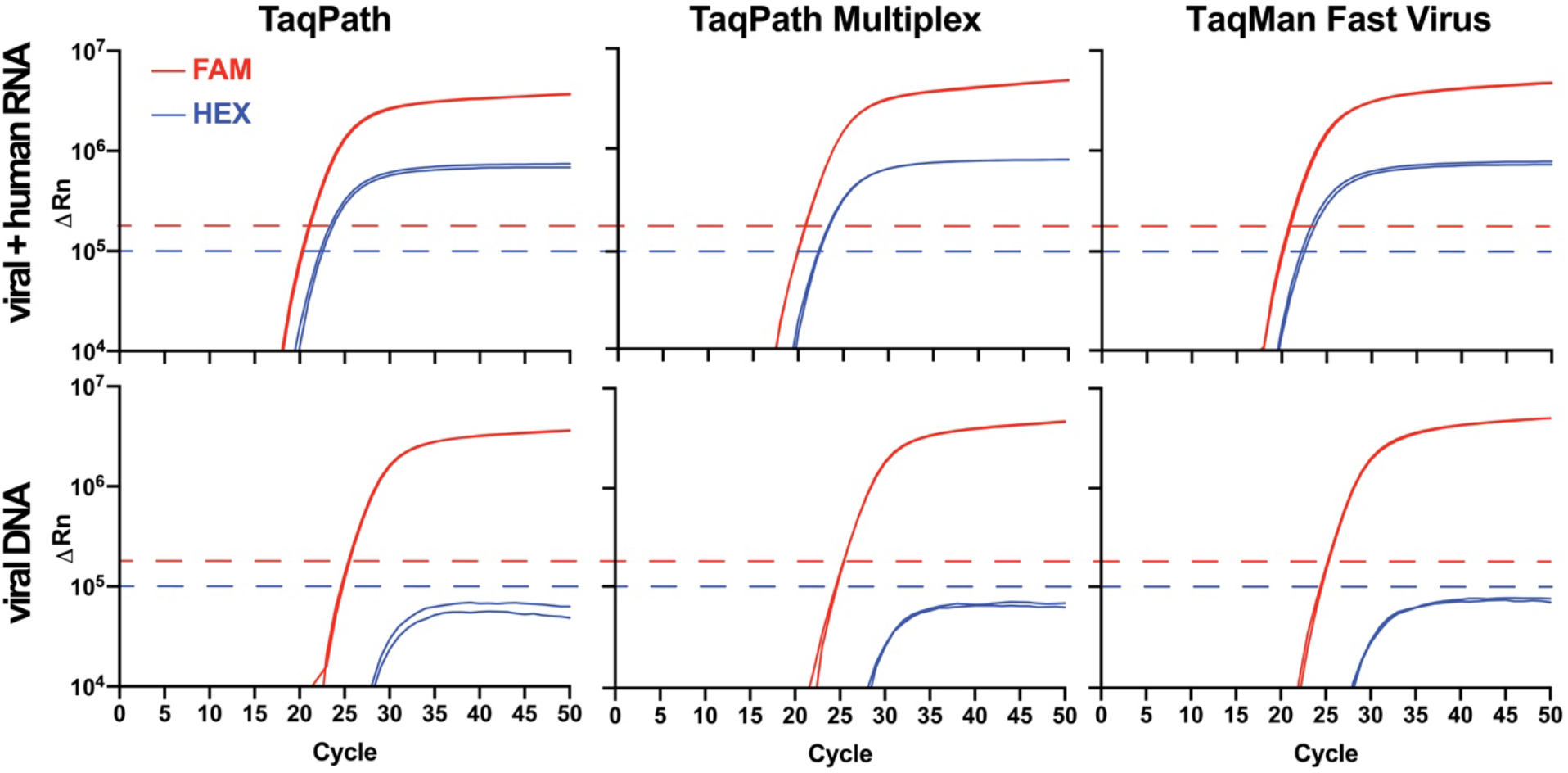
Performance of Georgia Tech multiplex primers and probes in several commercially available master mixes. GT multiplex primer/probe performance in commercial TaqPath, TaqPath Multiplex, and TaqMan Fast Virus 1-Step master mixes. Commercial master mix identity had no detectable impact on performance of the GT-made multiplex primer/probe mix. Due to the proximity of FAM and HEX channels, bleed-through from the FAM into the HEX channel was observed (see bottom nCov plasmid panels), but was of lower intensity than signal generated by the HEX-RP-BHQ1 probe (see top panels) and did not interfere with analyses when the HEX fluorescence threshold (blue dashed line) was set above the bleed-through noise. Template in the top row consisted of synthetic SARS-CoV-2 RNA (ATCC) mixed with HEK293T RNA. Results are consistent with those expected for a positive patient sample. A negative sample would consist of a single amplification curve in the HEX channel (blue line). Template in the bottom row was 2019_nCoV_N_Positive Control (IDT) plasmid DNA. Results are plotted logarithmically.

Consistent with the greater quantum yield of FAM relative to HEX, probes that contained FAM (λ_abs_ =494/λ_em_=518nm) generated a greater ΔRn signal than did probes that contained HEX (λ_abs_=535/λ_em_=556nm) in RT-qPCR experiments (**Supp. Fig. S2C**). Rn is the normalized reporter fluorescent dye signal normalized to the passive reference dye, and ΔRn is Rn value of the experimental sample minus the instrument baseline signal. In fact, the spectral characteristics and intensity of the FAM signal were such that a portion of the signal could be observed in the adjacent HEX channel of the instrument, albeit at a much lower intensity (‘bleed-through’; **Fig. 2**). The observed bleed-through of the FAM signal into the HEX channel likely arises due to spectral overlap of both HEX and FAM with the broad blue LED excitation (470/40 nm) in the StepOnePlus and QuantStudio 6 Flex instruments. Since the maximum ΔRn of the true HEX signal generated by the RP probe was always greater than the ΔRn from the bleed-through, unambiguous detection of HEX-RP-BHQ1 signal, indicating the presence of RNase P RNA in the sample, was possible by setting the HEX channel threshold above the ΔRn plateau of the bleed-through noise (**Fig. 2**). To address this complication in a clinical setting, a simple MATLAB script was written to interpret the multiplex results in the context of bleed-through and provide a color-coded read out in Microsoft Excel (https://github.com/rmannino3/COVID19DataAnalysis).

### Protein and enzyme production

#### RT and RTX

We obtained plasmids for two RT enzymes, an MMLV RT containing six mutations (15) and RTX, an engineered xenopolymerase with proofreading activity (16,17). Both enzymes were purified to near homogeneity (**Supp. Fig. S3A, B**) at high yield (10 mg/L for GT-MMLV and 4 mg/L for GT-RTX) by Ni^2+^-affinity chromatography followed by a second polishing step. RT activity was tested with in-house primers and templates from other projects (**Supp. Fig. S3A**) and remained highly active in RT-qPCR for ~2.5 months; longer term storage may require further optimization of the storage conditions. Characterization by OMNISEC reveals that GT-RTX is a dimer (167 kDa, **Supp. Fig. S3B**), compared to the monomeric MMLV RT (18). In line with Bhadra *et al*.(16), GT-RTX showed strong performance in RT-qPCR in the presence of SUPERase•In, a DTT-independent commercial RNase A inhibitor cocktail. However, when the DTT required for RNaseOUT was added, the ΔRn plateau for the reaction was low (**Supp. Fig. S3B**). Our priority was reliance on components that could be manufactured at GT, and since production of our DTT-dependent GT-rRI was successful (see Ribonuclease Inhibitor section), we did not further pursue use of GT-RTX in our master mix.

#### *Taq* polymerase

We considered five *Taq* constructs (Table 2) and two hot-start options. The best yields were obtained when T7-inducible *Taq* plasmids were transformed and grown in *E. coli* ArcticExpress with Superior Broth or in *E. coli* BL21 (DE3) using autoinduction medium or 2xYT broth. Still, expression yields (~0.5 mg/L culture at best) were notably less than the other proteins produced as part of this project.

**Table 2.** *Taq* polymerases tested in this project.

| Plasmid description | Antibiotic resistance | Expression *E. coli* strain |
| --- | --- | --- |
| pACYC | Chloramphenicol | HB101 |
| pAKTaq | Ampicillin | BL21 (DE3) |
| Nterm His *Taq* in pET28a | Kanamycin | BL21 (DE3), ArcticExpress |
| Cterm His *Taq* in pET20b | Ampicillin | BL21 (DE3), ArcticExpress |
| Sso7d-*Taq* in pET20b | Ampicillin | BL21 (DE3), ArcticExpress |

GT-*Taq* lacking affinity tags was used in our initial RT-qPCR formulations (see below). Of the purification strategies tested, a short heating step followed by anion exchange column chromatography, similar to that described by Desai and Pfaffle (19) was the most robust, reproducible and practical method. To save time and resources we conducted buffer exchange by using centrifugal devices or a PD-10 column, rather than standard dialysis. The purity of this *Taq* was less than other preparations we tested (Supp. Fig. S4A), e.g. that published by Engelke *et al*. involving polyethylenimine precipitation and weak cation exchange (not shown) (20), but this did not negatively impact enzyme performance. *GT-Taq* was highly active after storage for over 2 months.

GT-His-*Taq*, with a WT sequence and an N-terminal hexahistidine tag and purified only by Ni^2+^-affinity chromatography (Supp. Fig. S4A), was used in the final RT-qPCR formulation (see below). Addition of the hexahistidine tag streamlined protocols by enabling a purification scheme similar to that of GT-MMLV (above) and GT-rRI (below). C-terminally His-tagged *Taq* polymerase expressed at too low a level to warrant further consideration (not shown). Despite concerns about the possibility of protein contaminants or residual genomic DNA after purification, GT-His-*Taq* did not appear to benefit from a final anion exchange step (see Experimental Procedures, Supp. Fig. S4A). If residual genomic DNA is present in our purified polymerase, it apparently does not interfere with probe detection of viral amplicons in RT-qPCR. GT-His-*Taq* purified in one step was active in PCR (Supp. Fig. S4A) and exhibited robust enzyme activity in RT-qPCR (see below) for at least 2 months, after which point the supply was depleted from use in experiments.

In addition to GT-His-Taq, we considered the ssod7-*Taq* chimera, a more efficient *Taq* polymerase compared to WT *Taq* (21). Ssod7-Taq (**Supp. Fig. S4A**) expressed in significantly higher yield (2 mg/L) than any WT *Taq* polymerases we tested. Although sso7d-*Taq* performed as well as WT *Taq* in PCR and RT-qPCR (not shown), we were unable to identify conditions for storage of this enzyme. This version of *Taq* shows great promise but would only have full utility once storage issues are resolved.

Finally, our attempts to evaluate and develop hot-start technology (22) merit discussion. Hot-start is intended to minimize primer-dimer formation and premature extension of PCR products during reaction assembly and reverse transcription (23) by inhibiting *Taq* polymerase at low temperatures. We evaluated a commercial hot-start antibody (Supp. Fig. S4B) alongside two alternative hot-start approaches produced in-house: *Taq* mutant I705L (24) (**Supp. Fig. S4C**), and a variety of DNA aptamers from the literature (**Supp. Table S1**). Consistent with literature reports, the commercial hot-start antibody, the I705L *Taq* variant and some DNA aptamers did inhibit Taq polymerase at room temperature. However, only the hot-start antibody and, to a lesser extent aptamer-based One*Taq*® Hot Start DNA Polymerase, inhibited *Taq* at 37°C or 50°C. Hot start technologies tested here by RT-qPCR did not noticeably improve threshold cycle (Ct) values or fluorescence signals (data not shown).

Ribonuclease inhibitor. Though we did not detect RNase contamination in the purified GT enzymes we tested (**Supp. Fig. S5A**), RNase activity is anticipated in human and environmental samples. Mammalian RNase A is inhibited by RI, a leucine-rich repeat protein (25) bearing numerous reduced Cys residues. Inhibition of disulfide bond formation by a reducing agent such as DTT is particularly challenging with RI because it contains two pairs of adjacent Cys residues (25,26). We focused on porcine RI, which is known to be an effective RNase inhibitor and is amenable to recombinant production (27). Our GT-rRI has an N-terminal His-tag because that variant expressed better (1.5 mg/L) than the C-terminal His-tag. However, contrary to prior reports (28,29), the presence of DTT in the media did not alter our yield (not shown). Purification was carried out in the presence of fresh DTT. GT-rRI eluted from the Ni^2+^-affinity column at a high level of purity (**Supp. Fig. S5B**) and did not require further column purification. Inhibition of RNase A by GT-rRI was comparable to that by commercial RNaseOUT (**Supp. Fig. S5B**). GT-rRI was stored in aliquots of ~1 mg/mL in the presence of 8 mM DTT and was used in RT-qPCRs at volumes of 0.5-1 μL/20 μL (see below). No detectable change in inhibitor performance was observed over a two-month period.

### Formulation

Our goal was to develop an RT-qPCR master mix that, combined with our own primers and probes, would match the performance of commercial alternatives and would tolerate long-term storage. Commercial 1-step mixes contain proprietary additives for storage and improved performance. In early experiments we evaluated RT-qPCR formulations using agarose gel-based analysis. However, the size of the N1 amplicon is nearly identical to that of the primer dimer (**Supp. Fig. S3A**), complicating interpretation by that method. We found RT-qPCR to be a more direct route to feedback on formulation. For RT-qPCR described here we used Georgia Tech cycling conditions listed in **Table 3**.

**Table 3.** RT-qPCR thermal cycling conditions.

| Step | CDC(55) | Georgia Tech |
| --- | --- | --- |
| (1) UNG incubation | 25°C, 2 min | ---- |
| (2) Reverse transcription | 50°C, 15 min | 50°C, 15 min |
| (3) RT inactivation and/or DNA polymerase activation | 95°C, 2 min | 95°C, 5 min |
| (4) Denaturation | 95°C, 3 s | 95°C, 15 s |
| (5) Annealing and extension (fluorescence collection) | 55°C, 30 s | 55°C, 30 s |
| Number cycles of steps 4 and 5 | 45 | 45 |

RT and *Taq* share substrate dNTPs and cofactor Mg^2+^ cations but perform optimally under distinct reaction conditions. Further, RT can inhibit *Taq* (30). Therefore, any given formulation must be a compromise. Commercial RT buffers are at lower pH and higher salt (predominantly KCl) and Mg^2+^ than *Taq* buffers. Commercial *Taq* buffers also often contain low levels of detergent (Tween-20, Nonidet P-40, Triton X-100). Although we experimentally varied pH and ionic strength during our optimizations, our starting RT-qPCR buffer composition (50 mM Tris pH 8.3, 75 mM KCl, 3 mM MgCl_2_, 5 mM DTT, based on the buffer used with SuperScript III RT) yielded a fluorescence signal similar to TaqPath and could not be improved upon (**Table 4, Fig. 3**). RT-qPCR fluorescence was sensitive to the concentration of *Taq* in the reaction, but not of the RT (**Supp. Fig. S6**). Of the various *GT-Taq* enzymes tested, both GT*-Taq* (no tags) and GT-His*-Taq* performed well as long as the concentration was at least ~0.1 mg/mL in the final storage buffer. The addition of modest concentrations of salts such as (NH_4_hSO_4_ (not shown), CHAPSO (**Fig. 3A**), or other detergents (31) (not shown) to master mix containing GT*-Taq* enzymes resulted in the reduction of RT-qPCR fluorescence signal. Interestingly, initial optimization with Platinum II Hot-start *Taq* (**Supp. Fig. S7; Supp. Table S2**), which we tested in some early formulations, did not translate to the same optimal conditions for GT-*Taq*. Thus, in addition to the presence of the hot-start antibody, there may be other differences between the WT *Taq* we produced and Platinum II *Taq*. Finally, the addition of ROX^TM^ reference dye improved instrument baseline values across all experiments (not shown).

**Figure 3.**
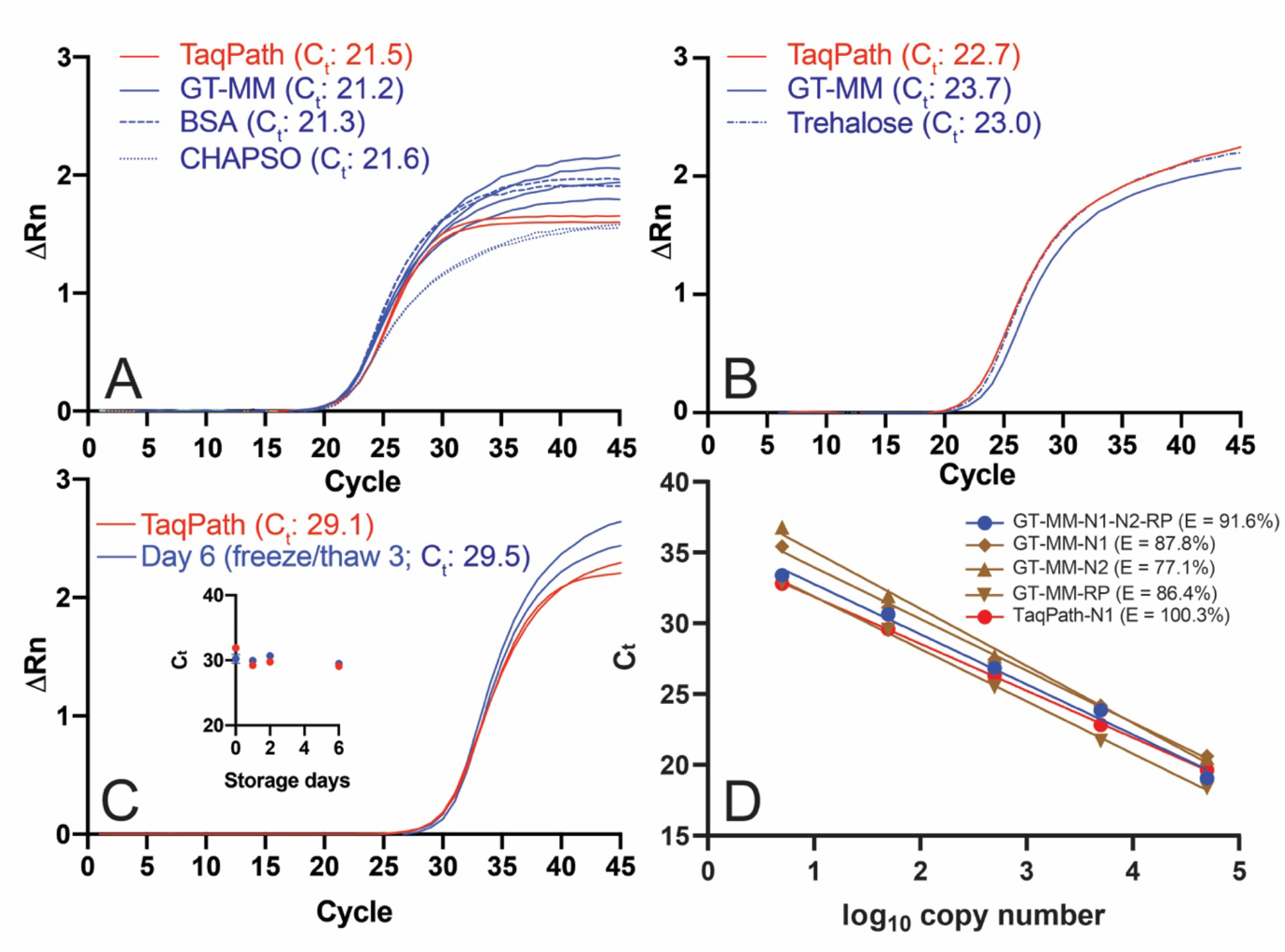
Performance of GT RT-qPCR master mix. RT-qPCRs were performed with the Georgia Tech thermal cycling conditions (**Table 2**) and GT-Master Mix components (**Table 4**) with ATCC synthetic viral RNA (ATCC® VR-3276SD™) and Ct values determined using a threshold of 0.1 unless otherwise noted. (**A)** Effect of CHAPSO (0.1%) and BSA (0.5 mg/mL) on GT master mix performance with IDT N1 primers and 50,000 copies of synthetic viral RNA. The no template control did not amplify. (**B)** Performance of GT multiplex primers and probes with 50,000 copies viral RNA with GT-Master Mix, compared to TaqPath, and effect of trehalose (9.5%). (**C)** Performance of GT-Master Mix (MM) with IDT N1 primers and 500 copies of synthetic viral RNA, compared to TaqPath, after three freeze/thaw cycles (six days of storage) at 2x concentration. *Inset:* Ct for GT-Master Mix and TaqPath over six days of storage. (**D)** qPCR efficiency (E = 10 ^(−1/slope)^ −1) using auto threshold. GT-Master Mix and GT multiplex primers (N1 and N2 FAM readout, blue): 91.6%; GT-Master Mix and GT singleplex primers (brown): 87.8% for GT-N1 primer/probe (diamond), 77.1% GT-N2 primer/probe (triangle), 86.4% GT-RP primer/probe (inverted triangle); 100.3% TaqPath with GT-N1 primer/probe (red). Singleplex RT-qPCRs were performed with a mix of full-length viral RNA and HEK293T total RNA.

**Table 4.** GT RT-qPCR test kit formulation.

| **Component** | **Stock** | **Volume (μL)** | **Final concentration** |
| --- | --- | --- | --- |
| **Template** | Quantitative Synthetic SARS-CoV-2 RNA: ORF, E, N (ATCC® VR-3276SD™) | 0.5-5 | 5-50,000 copies |
|  | or |  |  |
|  | Full-length viral RNA + HEK293T total RNA | 5 | 5-50,000 copies viral RNA + 0.02-200 ng HEK293T total RNA |
| **Primer/probe:** | GT singleplex or multiplex primer/probe mix | 1.5 | 500 nM primers<br>125 nM probes |
| **GT-Master mix:** | 5X buffer:<br>250 mM Tris pH 8.3,<br>375 mM KCl,<br>15 mM MgCl<sub>2</sub>,<br>47.5% trehalose | 4.0 | 1X buffer:<br>50 mM Tris pH 8.3,<br>75 mM KCl,<br>3 mM MgCl<sub>2</sub>,<br>9.5% trehalose |
|  | 10 mM dNTP | 0.8 | 400 μM |
|  | 100 mM DTT | 1.0 | 5 mM |
|  | GT-rRI<sup>2</sup> | 1.0 | 50 μg/mL |
|  | GT-His-Taq<sup>2</sup> | 1.0 | 7.5 μg/mL |
|  | GT-MMLV<sup>2</sup> | 0.5 | 3.3 μg/mL |
|  | 20 mg/mL BSA | 1.0 | 1 mg/mL |
|  | 25 μM ROX | 0.4 | 500 nM |
| **Water:** | Molecular biology grade water | xx |  |
| **Total volume** |  | **20** |  |

<sup>1</sup>Can be prepared as a 2X mix and stored at -20°C.
<sup>2</sup>See Experimental Procedures. Concentration or volume should be adjusted for activity.

Having identified buffer and enzyme conditions that yield fluorescence curves similar to TaqPath (for the GT-master mix (hereafter, GT-MM; **Fig.3A**)), we next examined the effects of stabilizers that serve dual purposes for enzyme activity (32,33) and stability (34,35). Addition of known stabilizers BSA and trehalose to our master mix did not affect RT-qPCR performance (**Fig. 3A, Fig. 3B**) so both were retained in the final formulation (trehalose, 9.5%; BSA, 1 mg/mL; **Table 4**). Other additives such as betaine (0.5 or 1 M), a secondary structure reducer, or oligo (dT) and a random DNA hexamer, which enhance reverse transcription, did not improve Ct values or fluorescence signals (data not shown). Three freeze/thaw cycles and storage for six days at −20 °C did not affect performance our master mix (**Fig. 3C**) in the presence of stabilizers. Finally, the qPCR efficiency over a 5-log range (5-50,000 copies RNA) in our GT-MM for the multiplex primer set was 91.6% (*r*^2^ = 0.991), similar to the efficiency observed when the singleplex primers and probes were tested; 87.8% (*r*^2^=0.997) for the GT-N1 primer and probe, 77.1% (*r*^2^=0.995) for the GT-N2 probe, and 86.4% (*r*^2^=0.999) for the GT-RP probe (**Fig. 3D**). For comparison, efficiency of the GT-N1 primer and probe in TaqPath was 100.3% (*r*^2^=0.999) (**Fig. 3D**). The GT multiplex assay is linear over the same 5-log range as TaqPath suggesting a similar limit of detection (**Fig. 3D**). In sum, our GT RT-qPCR assay, composed of proteins and enzymes produced in house, with either singleplex or multiplexed primers and probes kit exhibits a high level of qPCR efficiency and storage stability.

### Environmental testing

We developed a straightforward protocol (**Fig. 4A**) to monitor for the presence of viral DNA or RNA in our laboratory spaces. Our method includes the collection, preservation, and quantification of viral DNA or RNA by RT-qPCR. The procedure does not require an RNA extraction step (36-38). As the environmental testing aspect of the overall project was established in parallel with in-house RT-qPCR assay kit development, commercial master mix, primers and probes were used in environmental survey RT-qPCR. A high level of efficiency and sensitivity were established for both DNA and RNA samples (**Fig. 4B**).

**Figure 4.**
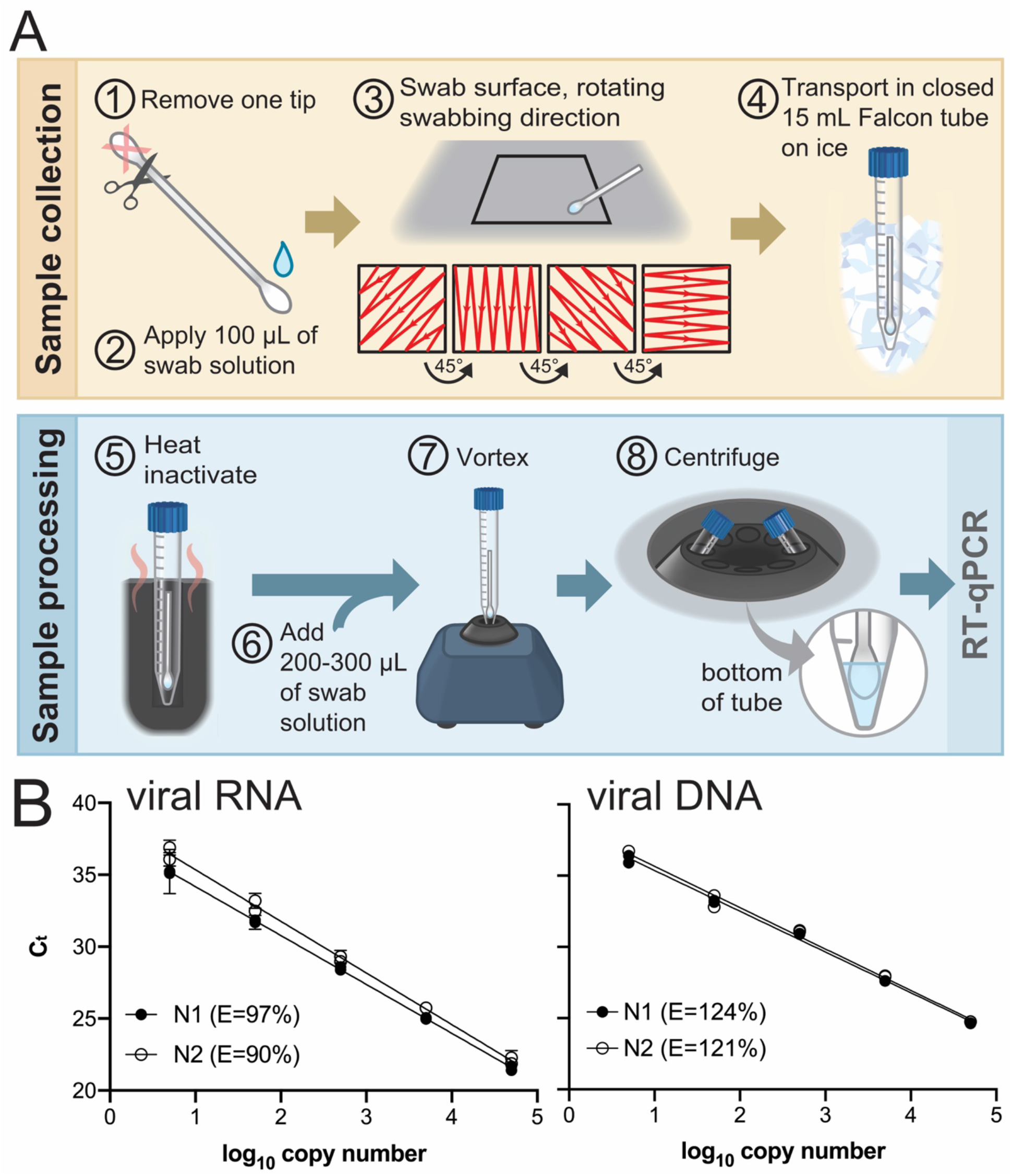
Environmental testing protocol and qPCR standard curve. **(A)** Environmental testing protocol (see text). (**B)** Standard curves used to calculate the magnitude of environmental surface contamination and qPCR efficiencies (E = 10 ^(−1/slope)^ −1) using TaqPath and IDT CDC primers and probes. Left template, quantitative synthetic SARS-CoV-2 RNA (ATCC #VR-3276SD), N1 *r*^2^=0.9933, N2 *r*^2^=0.9936. Right template, positive control plasmid viral DNA (IDT #10006625), N1 *r*^2^= 0.9965, N2 *r*^2^=0.9917.

Swab material and sample collection methods were evaluated during protocol development, as these are known to be important for downstream detection (39,40). Although liquid recovery was slightly better when knitted polyester swabs were used, lightly moistened cotton swabs were cheap, available, effective and reliable (**Supp. Fig. S8A**). We identified 0.5% Triton X-100 with 0.05 mM EDTA as a suitable medium for wetting the swab prior to sample collection, lysing the virus, and protecting viral RNA from degradation; it did not interfere with downstream RT-qPCR (**Supp. Fig. S8B**). The preservation of RNA is likely due to EDTA chelation of metal ions, which inhibits metal-mediated cleavage of RNA (41,42). Heating the sample prior to opening the container ensured complete lysis of viral particles (**Supp. Fig. S8C**), and likely contributed to the high level of RNA recovery from the swab.

In addition to monitoring the presence of RNA on surfaces throughout our campus buildings and laboratories, environmental testing was used to address the issue of cross-contamination of our probes and primers with the DNA template used as a positive control in the qPCR assay (see Primers and Probes above). Environmental testing detected the SARS-CoV-2 positive control plasmid DNA on surfaces and equipment in the laboratory in which the plasmid was handled, as well as in remote laboratories, indicating that it had been transferred, likely by personnel movements (**Supp. Fig. S8D**). The extreme sensitivity of RT-qPCR—the presence of even a single plasmid copy may give rise to an amplification signal—demands a level of stringency that is unfamiliar to most academic biochemists. To combat the contamination, we treated surfaces and pipettes with 10% bleach which was shown by subsequent rounds of environmental testing to be an effective means of eliminating plasmid DNA. After new primers and probes were synthesized, all RT-qPCR components including enzymes and buffers were tested exhaustively (to ensure the absence of contaminating viral template. The DNA plasmid was isolated to one laboratory, separate from other assay components. Long term, the best practice is likely off-site maintenance of positive control plasmids.

## Discussion

At the start of the COVID-19 pandemic, supply chain instability delayed testing for SARS-CoV-2 infection, particularly in the United States (43). Even though the supply chain is more stable now, uncertainty exists about sustained access to testing (44). At our large, residential state university in a major metropolitan area with a rapidly rising case rate, we anticipated a need for increased testing and monitoring through the fall semester and beyond.

In three months, we formulated a functional SARS-CoV-2 assay that compares favorably to commercially available RT-qPCR kits. Our assay comprises a master mix as well as primers and probes identical to validated CDC sequences. Initially, the CDC was the sole source of primers and probes to CLIA labs, but upon discovery of contamination issues (11), the oligomers from commercial labs were approved for use. Allowing external suppliers, including academic labs, to supply primers and probes to CLIA labs has bolstered supply availability.

In GT-MM, the efficiencies of our GT primer and probe sequences met (multiplex) or closely approached (singleplex) our efficiency target of 90-110% with high linearity (*r*^2^>0.990), indicating minimal primer dimers or non-specific amplification (45). The efficiency and linearity of our multiplex kit over a five-log concentration range is competitive with other kits that have received EUA for SARS-CoV-2 testing. A full ‘bridging study’ of the multiplex kit with lower limit of detection and clinically relevant samples is planned.

GT-MM is composed of affinity-purified GT-rRI, GT-His-*Taq*, and GT-MMLV at defined concentrations, a compatible buffer containing cationic cofactors, plus BSA and trehalose for stability and long-term storage. Even though RTX (5), a single enzyme with both RT and DNA polymerase activities that we considered for inclusion in GT-MM, was incompatible with the DTT required to stabilize GT-rRI, the ability of RTX to amplify the target indicates that it may prove useful in other contexts. Notably, our formulation does not include a hot start *Taq*; we found that performance of commercial hot start *Taq* depended on the buffer used, and under the final buffer conditions selected for the GT-Master Mix, our non-hot start GT-His-Taq outperformed hot start *Taq* in RT-qPCR.

Our protocol to test surfaces for SARS-CoV-2 RNA will be useful for monitoring viral deposition in the environment. While the presence of viral RNA on surfaces does not necessarily indicate live virus or suggest a source of viral transmission, monitoring high touch surfaces on a residential college campus will be beneficial to evaluate the effectiveness of preventative decontamination protocols. We found that a minimum swab medium consisting of water and EDTA is sufficient for viral recovery from surfaces and prevention of RNA degradation. While the inclusion of detergents disrupts the viral envelope, heat was most important to ensure viral lysis. To further improve the detection limit of our environmental protocol, additional reduction of volume may be required. Addition of proteinase K or RNase inhibitors to the swab medium may enable sample transport at room temperature.

One advantage of our environmental sampling method is that we minimized the volume of liquid used in all steps of the process, which maintains high sample concentration and negates the need for a separate RNA extraction step. Dry or nominally wet swabs (39,46,47) used here are an attractive replacement for VTM, a viral culturing medium used routinely during clinical SARS-CoV-2 sample collection and transport. VTM increases exposure risks during collection, transport, and handling of live virus and introduces large quantities (3 mL) of biological material. Such solutions are prone to spillage during transport (personal communication), dilute the swab sample by at least 100-fold, and must be removed through an extraction protocol before RT-qPCR can be conducted. Our wet swabs are also simpler than non-biological commercial substitutes such as DNA/RNA Shield (Zymo Research) or Trizol (Thermo Fisher).

The goal of the Georgia Tech COVID-19 Test Kit Support Group was to create contingency SARS-CoV-2 diagnostic test components in the face of supply line insecurity. We translated published information about RT-qPCR and sophisticated commercial kits into a series of fundamental protocols, executable with consumables and equipment routinely used in academic biochemistry laboratories. While most assay reagents were produced in-house, key specialty chemicals still needed to be purchased (phosphoramidites and fluorophores used in primer and probe synthesis; dNTPs, ROX^™^, molecular biology grade DTT, BSA, and trehalose used in the GT-MM). Our blueprint should be readily reproducible by research teams at other academic institutions, and our protocols may be modified and adapted to enable SARS-CoV-2 detection in more resource-limited settings. With a detailed protocol for an RT-qPCR assay in hand, we can maintain a pipeline for kit production and file for an EUA in order to backfill master mix and primers should new shortages arise. We can also monitor the presence of SARS-CoV-2 on high-touch surfaces throughout our campus community. In the long term, our protocols should be adaptable to the detection of other novel or seasonal infectious viral agents.

## Experimental Procedures

### Primers and probes

#### Synthesis

Primers and probe oligonucleotides (**Table 1**) were synthesized using an ASM-2000 high throughput DNA/RNA synthesizer (Biosset). Primer oligonucleotides were synthesized at 50-100 nmol scale with the 1000 Å universal control pore glass (CPG) support (Glen Research). Dual-labeled fluorophore/quencher probes were synthesized at 50-100 nmol scale with 4’-(2-Nitro-4-toluyldiazo)-2’-methoxy-5’-methyl-azobenzene-4”-(N-ethyl-2-O-(4,4’-dimethoxytrityl))-N-ethyl-2-O-glycolate-CPG (3’-BHQ1-CPG, Glen Research). The incubation time for coupling the fluorophore to the 5’-terminus was extended from manufacturer recommendations to ensure high efficiency; the 5’-hexachloro-fluorescein phosphoramidite (HEX) was coupled for 4.5 min and the fluorescein phosphoramidite (FAM) was coupled for 18 min. The coupling efficiency of the synthesis was monitored by orange color trityl fractions from the deprotection steps. The 4,4’-dimethoxytrityl (DMT) group on the 5’-FAM phosphoramidite was not cleaved off for downstream cartridge purification, while the HEX (no DMT protection) probe synthesis was completed the same as primers. The synthesized oligonucleotide primers were cleaved from the CPG support by 4x treatment with 200 μL 30% ammonium hydroxide for 20 min (800 μL final volume). The cleaved oligonucleotide primers in ammonium hydroxide were deprotected at 45°C for ~15 h and then vacuum dried under low heat (40-45°C) for ~4 h. The pellets were rehydrated with 1X IDTE buffer pH 7.5 (IDT) to a stock concentration of 6.7 μM. The same cleavage protocol was used for the probes. The cleaved FAM probe product was column purified using a Glen-Pak™ DNA purification cartridge. The eluted FAM probe was vacuum dried and resuspended in 1X IDTE buffer pH 7.5 (IDT) to a stock concentration of 1.7 μM. Concentrations were determined using a DeNovix DS-11 FX Spectrophotometer (1 OD_260_ = 33 ng/μL for ssDNA) and then converted to molar concentration using the respective molecular weight. The deprotected HEX probe was dried the same as the primers and used without further purification (stock concentration = 1.7 μM).

### Quality control

The purity of probes (3 μM) was evaluated by analytical HPLC performed at 20°C on an Agilent 1260 Infinity Series HPLC with a Kinetex XB-C18 column (Phenomenex, 2.6 μm, 150×2.1 mm). Buffer “A” was composed of 0.1 M ammonium acetate in water (pH 6.7) and Buffer “B” was composed of 0.1 M ammonium acetate in 50% acetonitrile. Analytical HPLC was run with a flow rate of 0.30 mL/min by running 30% buffer “B” for 5 min, then a gradient of 30-60% buffer “B” for 25 min, then finally 30% buffer “B” for another 5 min.

For quality control, 10 pmol of each primer was 5’ end-labeled with 100 pmol of ^32^P-ATP (Perkin Elmer) at 37°C for 30 min using T4 polynucleotide kinase (NEB #M0201). The enzyme was inactivated by incubation at 65°C for 20 min. Labeled oligonucleotides were run on a denaturing 20% polyacrylamide gel at 14 W for 60 min. The gel was exposed to a phosphor screen and scanned on a Typhoon FLA 9500 (GE Healthcare).

Performance of in-house primers and probes was validated by RT-qPCR using a mixture of TaqPath 1-Step Master Mix (5 μL/rxn), Georgia Tech singleplex primers/probe or multiplex primers/probes (1.5 μL/rxn), and nuclease-free water (8.5 μL/rxn). A panel of reactions was performed in which various templates were used: synthetic SARS-CoV-2 RNA (ATCC #VR-3276SD), HEK293T cell RNA (containing RNase P RNA) generated in-house by TRIzol (Invitrogen #15596026) extraction of HEK293T cells grown to 60% confluency, a mixture of SARS-CoV-2 RNA and HEK293T cell RNA, nuclease-free water (negative, no template control), or SARS-CoV-2 plasmid (2019-nCoV_N_Positive Control, IDT #10006625, 10,000-50,000 copies/rxn). To determine compatibility between the Georgia Tech multiplex primers/probes mix and commercial master mixes, TaqPath 1-Step Master Mix, TaqPath 1-Step Multiplex Master Mix, and TaqMan Fast Virus 1-Step Master Mix (all ThermoFisher Scientific) were prepared according to manufacturer instructions and templates were either nuclease-free water (negative, no-template control), a mixture of SARS-CoV-2 RNA and HEK293T cell RNA, or SARS-CoV-2 plasmid (positive control). RT-qPCR cycling conditions were used as listed for CDC but omitting the UNG step (**Table 3**).To test for contamination, nuclease-free water (7 μL/rxn), plus IDT N1 primers/probe mix (1.5 μL/rxn), IDT N2 primers/probe mix (1.5 μL/rxn), were added in turn to TaqPath (5 uL/rxn) with the newly synthesized Georgia Tech primer or probe added as the template (5 μL/rxn); the Georgia Tech primers were tested at a final concentration of 500 nM and Georgia Tech probes were tested at a final concentration of 125 nM. IDT SARS-CoV-2 plasmid served as the positive control. After cycling in either a StepOnePlus (Applied BioSystems) or QuantStudio 6 Flex (Thermo Fisher Scientific) qPCR instrument, results were evaluated to ensure that amplification occurred only in the expected samples/channels. The lack of amplification using IDT primers and probes, paired with amplification in the SARS-CoV-2 plasmid positive control, confirmed newly synthesized probes and primers were free of contamination.

### Multiplex assay interpretation

A MATLAB script (https://github.com/rmannino3/COVID19DataAnalysis) was written to interpret the results of RT-qPCR assays conducted with GT multiplexed (FAM/HEX) primers and probes based on 96-well data exported from QuantStudio 6 and StepOne Plus qPCR instruments. The user sets the cycle threshold (C_t_), as well as the location of both the positive and negative controls via an interactive prompt. A folder of Excel files containing RT-qPCR data (exported directly from the instrument) are processed to yield a visual readout. The output is a color-coded 96-well grid, with green corresponding to a Negative “-” result (no amplification in the FAM channel below the Ct *and* amplification in the HEX channel below the Ct), red corresponding to a Positive “+” result (amplification in the FAM channel below the C_t_ *and* amplification in the HEX channel below the C_t_), and yellow corresponding to an “Inconclusive” result (no amplification in the HEX channel below the C_t_). Finally, the script determines whether the HEX intensity threshold is set properly. In practice, the HEX intensity threshold should be set above the max intensity value of the HEX signal in the nCoV positive control wells, where no human RNA is present. The function provides a warning to the user in the MATLAB command window if this condition is not met.

### Expression and purification of GT-MMLV

The MMLV RT plasmid for the production of GT-MMLV was a kind gift from Dr. Amy Lee (Brandeis University). This MMLV RT lacks the RNase H activity of native RT, contains mutations that increase thermostability (15), and has been demonstrated effective in RT-qPCR (48). Sequencing (Eton Biosciences) confirmed the E69K, E302R, W313F, L435G, N454K, and D524N mutations (see **Supp. data files**). The plasmid was used to transform *E. coli* BL21 (DE3) or *E. coli* ArcticExpress (Agilent) by heat shock.

For growth with *E. coli* BL21 (DE3), a single colony was used to inoculate 25 mL of 2xYT Medium Broth (VWR) supplemented with 50 μg/mL kanamycin. The overnight culture was grown at 37°C for 13-14 h, shaking at 180 rpm. Ten milliliters of the overnight culture were used to inoculate two separate 2.8 L baffled flasks, each containing 1 L of 2xYT media supplemented with 50 μg/mL kanamycin. Cultures were grown at 30°C, with shaking at 180 rpm, until the OD_600_ reached 0.4, at which point the temperature was decreased to 16°C. After one additional hour, protein production was induced with 0.5 mM isopropyl-*β*-d-1 –thiogalactopyranoside (IPTG).Cultures were harvested 18 h post-induction by centrifugation (5,000 × g, 20 min, 4°C).

For purification of GT-MMLV from *E. coli* BL21 (DE3), the cell pellet was resuspended in 60 mL lysis buffer (**Supp. Table S3A**) supplemented with protease inhibitor (Pierce, EDTA-free). Cells were lysed on ice for 25 min with a QSonica CL-18 sonicator operating at 40% power (15 s on, 45 s off). Lysate was centrifuged (40,000 x g, 40 min, 4°C). The supernatant was loaded onto an AKTA Prime FPLC system equipped with a 5-mL HisTrap column (GE Healthcare) equilibrated with Buffer A (**Supp. Table S4A**), at a flow rate of 1 mL/min. The column was washed with 60 mL of Buffer A at 2 mL/min. GT-MMLV was eluted using a linear gradient to 100% Buffer B (**Supp. Table S4A**) over 40 mL. The target protein began eluting around 250 mM imidazole (~80% Buffer B; **Supp. Table S4A**). The purity of each fraction was evaluated by denaturing SDS-PAGE and the purest fractions were pooled. Approximately half of the pooled material was dialyzed overnight (~16-18 h) at 4°C using a 10,000 MWCO dialysis tubing against 2 L of 20 mM Tris-HCl (pH 7.5), 100 mM NaCl, 1 mM Tris (2-carboxyethyl)phosphine hydrochloride (TCEP-HCl), 0.01% nonyl phenoxypolyethoxylethanol (NP)-40, 10% glycerol (v/v). The protein was stored at −20°C after exchange with storage buffer (**Supp. Table S5A**) using a PD-10 size exclusion chromatography column (GE Healthcare).

The remaining pooled samples of GT-MMLV from *E. coli* BL21 (DE3) were subjected to further purification by anion exchange as follows: pooled fractions were dialyzed for 16 h at 4°C using 10,000 MWCO dialysis tubing against 2 L of dialysis buffer (20 mM Tris-HCl (pH 8.9), 50 mM KCl, 1 mM DTT, 10% glycerol (v/v)). The dialyzed protein was loaded at 1 mL/min into a 5-mL Hi-Trap Q column using AKTA Prime FPLC system pre-equilibrated with Buffer A (Supp. Table S6A). The column was washed at 2 mL/min with 10 mL of Buffer A. GT-MMLV was eluted at ~700 mM NaCl using a linear gradient to 100% Buffer B (**Supp. Table S6A**) over 40 mL. After SDS-PAGE analysis of elution fractions, the single purest fraction was buffer exchanged into storage buffer as above (**Supp. Table S5A**) using a 5 mL PD-10 desalting column, and stored in ~500 μL aliquots at −20°C. Protein concentration (2.6 mg/mL, ~9 mg total) was calculated using the Bradford method, but is likely overestimated due to the detergent in the storage buffer. Activity of both Ni-NTA purified and IEX-purified GT-MMLV were verified with RT-PCR using in-house protocols (49). Briefly, in a two-step RT-PCR, total RNA isolated from seaweed *Asparagopsis taxiformis* was used as template to amplify a 150-bp region of the gene encoding actin and visualized by agarose gel electrophoresis.

For expression of GT-MMLV in *E. coli* ArcticExpress (Agilent) described below, a single colony was used to inoculate 5 mL of Lysogeny broth (LB) supplemented with 25 μg/mL kanamycin (Sigma-Aldrich) and 10 μg/mL gentamicin (VWR) in a 50 mL Falcon tube, and incubated at 37°C overnight, shaking at 200 rpm. The next morning, 4 mL of the overnight culture was used to inoculate 200 mL of autoinduction media modified from Studier et al (50). Autoinduction media (1L) was prepared by combining a 960 ml solution composed of 100 ml of 10x phosphate buffered saline pH 7.4, 20 g tryptone, 5 g yeast extract, 5 g NaCl with 40 ml of a glucose mix (2 g lactose, 0.5 g glucose, 5 ml glycerol, 34 ml water). The 200 ml culture was incubated at 25°C, 150 rpm for 24 h. Cells were harvested by centrifugation (3500 x g, 20 min, 4°C). A total of 5.3 g cell mass was achieved (20 g/L). GT-MMLV in *E. coli* ArcticExpress was also expressed by adding 20 mL of inoculum to 2 L baffled flasks each containing 1 L Superior broth (US Biological) supplemented with 50 μg/mL kanamycin and 20 μg/mL gentamicin. These cultures were grown at 37 °C at 250 rpm until the OD600 reached 0.3 - 0.5, upon which the temperature of the incubator was dropped to 18°C. After ~1.5 h, IPTG was added to each flask at a final concentration of 1 mM and agitation was reduced to 200 rpm. Cultures were allowed to grow overnight for ~16-18 h post-induction before harvesting by centrifugation (5,000 × g, 10 min, 4°C). Cell pellets were flash cooled in liquid nitrogen and stored at −80°C.

For purification of GT-MMLV from *E. coli* ArcticExpress grown in autoinduction media, 2 g of cell mass was resuspended in 10 mL of lysis buffer (**Supp. Table S3A**) and sonicated on ice at 50% output = 10 W (Fisher Scientific Sonic Dismembrator) for 5 min. The slurry was centrifuged (16,000 × g, 30 min, 4°C), the supernatant was transferred to 1-mL Ni-NTA beads (MClabs) in a gravity column that had been equilibrated with Buffer A (**Supp. Table S4A**), and the slurry was incubated on ice for 30 min. After incubation, the column was allowed to drain using gravity and the Ni-NTA beads were washed with 10 column volumes (CV) Buffer A (**Supp. Table S4A**) and the RT was eluted with 2.5 mL of Buffer B (**Supp. Table S4A**). The GT-MMLV solution was immediately exchanged into 20 mM Tris pH 9.0, 1 mM DTT using a PD-10 column and the resulting 3.5 mL elution was purified using a 5 mL gravity Sepharose Q column (GE Healthcare) equilibrated with Buffer A (**Supp. Table S6A**). After draining the flow-through, the column was washed with 10 mL of 20 mM Tris pH 9.0, 1 mM DTT. The bound GT-MMLV eluted from the Q column with 10 mL of 20 mM Tris pH 9.0, 1 mM DTT, 300 mM KCl. A total of 2 mg GT-MMLV was recovered based on the Bradford method. The salt concentration was reduced to the 100 mM KCl using Macrosep concentrators and the final volume was adjusted to 0.065 mg/mL protein using storage buffer (Supp. Table S5A), dispensed into 50 μL aliquots, and stored at −20°C. A total of 30 mL of RT solution was achieved from the initial 2 g cell mass (~30,000 RT reactions for RT-qPCR).

To test GT-MMLV activity, a two-step endpoint RT-PCR was performed with N1 primers, SuperScript IV RT (Thermo Fisher Scientific #18090010) or GT-MMLV, and *GoTaq* (Promega #M3001). The template was 100,000 copies of Quantitative Synthetic SARS-CoV-2 RNA: ORF, E, N (ATCC® VR-3276SD™). The RT-PCR thermal cycling conditions were 55°C for 10 min, 95°C for 10 min, followed by 30 cycles of 95°C for 30 s, 55°C for 30 s, and 68°C for 30 s, with a final extension step at 68°C for 10 min. RT-PCR products were separated on a 2% agarose gel for 40 min at 160 V.

### Expression and purification of the xenopolymerase GT-RTX

The RTX plasmids (with (exo+) and without (exo-) exonuclease activity) (5,17) were a kind gift from Dr. Andrew Ellington (UT Austin). The RTX (exo+) plasmid (see **Supp. data files**) was used to transform *E. coli* BL21 (DE3) cells and a single colony was used to inoculate a 5 mL culture in LB supplemented with 100 ug/mL ampicillin, and was grown for 18 h at 37°C with shaking at 200 rpm. One mL of the culture was transferred to 50 mL of autoinduction media (see GT-MMLV above) supplemented with 100 μg/mL ampicillin and grown at 25°C, shaking at 150 rpm for 24 h. Cells were harvested by centrifugation (4,000 × g, 30 min, 4°C). The cell pellet (1 g) was resuspended in 5 mL lysis buffer (Supp. Table S3B) and sonicated at 50% output (Sonic Dismembrator, Fisher Scientific) for 4 min on ice. The lysate was cleared by centrifugation (4,000 × g, 30 min, 4°C). The supernatant was incubated in a thermomixer shaker (400 rpm, 10 min, 85°C) (Eppendorf) and centrifuged (16,000 × g, 20 min, 4°C). The supernatant was applied to a 1-mL Ni-NTA (MCLabs) gravity column equilibrated with Buffer A1 (**Supp. Table S4B**) and incubated on ice with intermittent inversion over the course of 30 min. After incubation the slurry was allowed to settle and the flow-through was discarded. The column was washed with 10 volumes of Buffer A1 (**Supp. Table S4B**) and 10 volumes of Buffer A2 (**Supp. Table S4B**). Bound protein was eluted with 2.5 mL of Buffer B (**Supp. Table S4B**).

For anion exchange using a PD-10 column, GT-RTX was exchanged into Buffer A (**Supp. Table S6B**) and the resulting 3.5 mL protein solution was applied onto a 5-mL Q-Sepharose (GE Healthcare) gravity column equilibrated with Buffer A (**Supp. Table S6B**). The column was washed with 5 volumes of Buffer A (**Supp. Table S6B**) and the GT-RTX was eluted with Buffer B (**Supp. Table S6B**). GT-RTX was concentrated using a Macrosep concentrator (10K MWCO), diluted into Buffer A (**Supp. Table S6B**), concentrated again, and diluted into 50% glycerol for final storage (Supp. Table S5B). GT-RTX (exo+) was tested for polymerase activity by PCR as described for *Taq* below. To analyze oligomeric state, GT-RTX was characterized by size exclusion chromatography equipped with OMNISEC REVEAL (Malvern), consisting of an analytical size exclusion column (Sepax Technologies, SRT SEC-300, 5-μm 300Å, 7.8×300 mm), a right (90°) and low (7°) angle SLS detector, an UV/Vis detector, a refractive index detector, and a viscometer.

### Expression and purification of *Taq* polymerases

Five *Taq* constructs were evaluated (**Table 2, Supp. Data files**). Plasmids for WT *Taq* polymerase lacking affinity tags were purchased from Addgene. To generate N-terminal hexahistidine tagged *Taq* polymerase (GT-His-*Taq)*, the pAKTaq plasmid (Addgene) and pET28a vector (Novagen) were cut with EcoR1 and Sal1 restriction enzymes and then ligated (Lucigen Rapid Ligation Kit) and transformed into *E. coli* strains for storage (DH5α) or expression (BL21 (DE3)). Plasmids for the fusion protein sso7d*-Taq* (21) and a *Taq* polymerase were cloned into the pET20b vector containing a C-terminal hexahistidine tag (Biobasic). The I705L mutation was introduced into GT-His*-Taq* by modified inverse PCR protocol, loosely based on a published method (51). Briefly, primers were designed to create an inverse PCR product with one primer harboring the mutation and the other primer blunt ending the mutation primer in the 3’ direction. A linear plasmid was created in exponential fashion which was then ligated according to the KLD Enzyme Mix protocol (NEB), where phosphokinase, ligase, and DpnI were mixed together to circularize the plasmid and digest the parent plasmid to only create colonies of mutational origin after transformation of the circularized plasmid. Plasmid fidelity was confirmed for each by DNA sequencing (Genscript or the Georgia Institute of Technology Molecular Biology Core Facility).

*Taq* polymerase lacking affinity tags was expressed and purified. After transformation of pACYC and pAKTaq into *E. coli HB101* and *E. coli* BL21 (DE3), respectively, cells were grown by a modified protocol from the literature(19) or in Superior broth with overnight cold induction as is often done for other challenging projects (52). For the former method, a single colony or stab from a glycerol or DMSO stock was used to inoculate 2 mL of 2xYT medium supplemented with the appropriate antibiotic (60 μg/mL ampicillin or 50 μg/mL kanamycin) in a 15 mL Falcon tube. After overnight (~16-18 h) growth at 37°C, shaking at 300 rpm, the cultures were pelleted (12,000 × g, 10 min, 4°C) and resuspended in 1 mL of fresh media with antibiotic. This culture was added to 2 L baffled flasks containing 250 mL 2xYT broth supplemented with the appropriate antibiotic for overnight (~16-18 h) growth at 37°C, shaking at 300 rpm. The following morning, 10 mL of the culture was pelleted (14,000 × g, 10 min, 4°C), for each 500 mL 2×YT to be cultured. Each pellet was resuspended in 1 mL 2×YT broth (with antibiotic) that had been removed from the 500 mL culture, and then added back to the bulk medium. Cells were monitored for growth by OD_600_ and induced with 1 mM IPTG upon reaching an OD_600_=0.3-0.4. Cells were grown overnight (~16-18 h) post induction at 37°C, shaking at 325 rpm and pelleted by centrifugation (14,000 x g, 10 min, 4°C) and either used directly in purification or preserved by flash-cooling in liquid nitrogen and stored at −80°C for future purification.

*Taq* polymerase lacking affinity tags was purified by a heating followed by anion exchange column chromatography. Buffer A (30 mL, Supp. Table S6C) supplemented with 4 mg/mL lysozyme was added to ~10 g thawed cell pellet, resuspended by inverting tubes, and incubated 15 min at room temperature to allow for lysis. This solution was combined with 30 mL of 2X storage buffer (40 mM Tris-HCl, pH 8.0, 100 mM KCl, 0.2 mM EDTA, 1% NP-40, 1% Tween 20, and 2 mM freshly prepared DTT; 1X buffer in **Supp. Table S5C**) and incubated for 15 min at room temperature. The 60 mL solution was then divided into two Falcon tubes and sonicated on ice at 65% power for ~5 min (2 s on, 2 s off) until the cells were lysed and the pellet was liquified. The two tubes were incubated in a 75°C water bath for 60 min with periodic gentle inversion. The lysate was centrifuged (13,000 x g, 30 min, 4°C), and the supernatant was filtered with a 0.44 μm syringe filter. The supernatant was loaded onto a 5 mL Hi-Trap Q FF column pre-equilibrated with Buffer A (**Supp. Table S6C**), then washed with 10-20 CV of Buffer A on an Akta Pure until the Abs_280_ returned to baseline. *Taq* was eluted with a 20 CV gradient to 100% Buffer B (**Supp. Table S6C**). As soon as the *Taq* peak eluted, samples were run on SDS-PAGE to evaluate purity. Fractions containing pure *Taq* were pooled and concentrated using an Amicon Ultra concentrator (30K MWCO). After the first concentration step, the Abs_280_ was measured and the concentration was estimated by the extinction coefficient (110,380 cm^-1^ M^-1^), with final concentration of 0.4 mg/mL. This sample was diluted into ~12 mL 2X storage buffer, concentrated again to <0.5 mL, and diluted 1:1 with sterile glycerol (**Supp. Table S5C**). After gentle mixing, the solutions were dispensed into twenty 50 μL aliquots, and stored at −20°C. Note that this method concentrates the detergent and therefore for subsequent purifications, the buffer exchange protocol was amended (see below).

His-tagged *Taq* constructs were expressed in *E. coli* BL21 (DE3) and *E. coli* ArcticExpress (Agilent) strains. Expression in BL21 (DE3) expression followed the protocol for GT-RTX except 200 mL autoinduction media (see GT-MMLV) supplemented with kanamycin (25 μg/mL) was used. Expression in *E. coli* ArcticExpress followed the same protocol as for GT-MMLV above. Comparable yields were achieved for all His-tagged *Taq* constructs (see Results).

For a two-step column purification protocol tested, 5 g of a GT-His-Taq cell pellet were resuspended with 25 mL of lysis buffer (**Supp. Table S3C**), supplemented with protease inhibitor cocktail (cOmplete Tablets EDTA-free, Roche)) and then lysed by sonication for a total of 5 min (20 s on, 20 s off) on ice. Cell debris was removed with ultracentrifugation (27,000 x g, 25 min, 4°C). The supernatant was heated at 75°C with intermittent inverting for 10 min and purified with a 1 mL HisTrap HP column. Briefly, the column was equilibrated with Buffer A (**Supp. Table S4C**), the sample was loaded and then washed with Buffer A until baseline absorbance was reestablished. GT-His-Taq was eluted using a linear gradient to 100% Buffer B (**Supp. Table S4C**) over 10 CV. Elution fractions were pooled and concentrated to ~0.5 mL using an Amicon Ultra concentrator (30K MWCO), then applied to a 5-mL HiTrap Q FF anion exchange column equilibrated with Buffer A (**Supp. Table S6D**). After loading the sample, the column was washed with Buffer A until baseline absorbance was reestablished, then a gradient to 100% Buffer B (**Supp. Table S6D**) over 10 CV was applied. After SDS-PAGE analysis of the elution peak, the cleanest fractions were pooled and concentrated to 0.5 mL, as before. This concentrated sample was diluted to 15 mL in 40 mM Tris-HCl pH 8.0, 100 mM KCl, 0.2 mM EDTA and concentrated again to ~0.5 mL. At this point, the concentration was measured by Abs_280_ as described above, yielding 0.1 mg/mL. Then, the sample was spiked with 1:10 dilution (v/v) of 10% NP-40, 10% Tween 20, 20 mM DTT. Finally, 50% glycerol was added to complete the storage buffer (**Supp. Table S5D**), the sample was mixed gently, and stored at −20°C in nineteen 50 μL aliquots.

For the GT-His*-Taq* that was ultimately used in the master mix (and for which the production and purification protocol yielded the most protein), the heating step above was omitted and the enzyme was purified solely by passage over a HisTrap column. Cells (5 g) were resuspended in lysis buffer and lysed by sonication as above. HisTrap purification proceeded as described above, and elution fractions were pooled and concentrated to 0.5 mL. The buffer was exchanged into 100 mM Tris pH 8.0 in an Amicon Ultra concentrator (30K MWCO) by diluting to ~12 mL and re-concentrating to 0.5 mL. After the concentration was determined to be 0.3 mg/mL by absorbance (280 nm, see above), the sample was diluted 1:1 (v/v) in sterile glycerol (**Supp. Table S5D**) and dispensed into twelve 50 μL aliquots for storage at −20°C. This quantity of enzyme translated to ~600 RT-qPCRs.

The I705L *Taq* polymerase variant and sso7d*-Taq* fusion protein were purified by the protocol established for GT-RTX purification (see above). After Q-Sepharose purification, the elution fraction was exchanged into 40 mM Tris pH 8.0, 100 mM KCl, 0.2 mM EDTA using a PD-10 column and concentrated in a Macrosep concentrator before dilution to a final buffer composition identical to that for the stringent protocol above (**Supp. Table S5D**), and stored at −20°C.

Polymerase activities for all *Taq* enzymes were tested using in-house primers and template generating a 1.2 kb fragment. PCR conditions were 98°C for 10 min followed by 45 cycles of 98°C for 10 s and 55°C for 60 s. The PCR product was purified and on a 1.2% agarose gel and visualized using the fluorescent Midori dye (VWR).

### Hot-start *Taq polymerase*

Candidate hot-start DNA aptamers identified from the literature (**Supp. Table S1**) were synthesized in the Georgia Tech Molecular Biology Core Facility using standard methodology. To test for hot-start activity, 10 pmoles of an 18-mer oligo (**Supp. Table S1**) was labeled with ^32^P-ATP and allowed to anneal to a 40-base oligo complimentary strand (**Supp. Table S1**). Aptamers (250 nM) were tested with a non-hot-start commercial DNA polymerase (OneTaq, optimized blend of *Taq* and Deep Vent DNA polymerase, NEB #M0480). The I705L *Taq* variant was compared to One*Taq*, as above, as well as an aptamer-based hot-start commercial DNA polymerase (Hot-Start OneTaq; NEB #M0481). For testing a commercial hot-start antibody, 1 U of non-hot-start *OneTaq* (#M0480) or standard *Taq* (NEB #M0273) were first mixed with 1 μL of Platinum *Taq* monoclonal antibody (Thermo Fisher Scientific #10965-028) on ice and incubated for 30 min. Samples were heated at 55°C for 30 s, and slowly cooled (0.2 °C s^-1^) to 20°C. The tubes were then placed on ice until addition to a ^32^P-ATP labeled mixture with sufficient dNTPs and 1X ThermoPol buffer (NEB #B9004) to ensure complete strand polymerization. The mixture was incubated for 15 min at 25, 37, and 50°C, or 95°C for 2 min and 75°C for 15 min, before the reaction was quenched using 95% formamide. A 20% polyacrylamide denaturing gel run at 14 W for 75 min was exposed to a phosphor screen and scanned on a Typhoon FLA 9500 (GE Healthcare).

### Expression and purification of porcine ribonuclease (RNase) inhibitor (GT-rRI)

Plasmids for recombinant ribonuclease inhibitor (rRI) containing an N-or C-terminal hexahistidine tag were purchased from Twist Biosciences. For large scale growth, 120 mL of *E. coli* ArcticExpress that had been transformed with the N-terminally tagged GT-rRI was divided into six 2-L baffled flasks, each containing 1 L of Superior Broth supplemented with 50 μg/mL kanamycin (Sigma-Aldrich) and 20 μg/mL gentamicin (VWR). Cultures were grown at 37°C, shaking at 250 rpm until an OD_600_ = 0.3-0.5 was reached, at which time the temperature of the incubator was dropped to 18°C. After ~1.5 h, IPTG was added to each flask at a final concentration of 1 mM and shaking was reduced to 200 rpm. Cultures were allowed to grow ~16-18 h overnight post-induction, then cells were harvested by centrifugation (5,000 × g, 10 min, 4°C). Pellets were flash cooled in liquid nitrogen and stored at −80°C.

GT-rRI was purified by first thawing and resuspending ~2.5 g cells in 12.5 mL of lysis buffer (**Supp. Table S3D**) supplemented with protease inhibitor cocktail (cOmplete Tablets EDTA-free, Roche). Resuspended cells were sonicated on ice at 50% power for 20 s on/off and inverted after each cycle for ~5 min until the viscosity decreased. Cell debris was pelleted by centrifugation (27,000 x g, 20 min, 4°C). The clarified lysate was loaded onto a 1 mL HisTrap column that had been equilibrated with Buffer A (**Supp. Table S4D**) and the lysate was washed with Buffer A until baseline was reached again. GT-rRI eluted during a gradient to 100% Buffer B (**Supp. Table S4D**) over 10 CVs. After analysis of elution fractions by SDS-PAGE, the purest fractions were pooled and concentrated to ~0.5 mL in Amicon Ultra concentrators (30K MWCO), diluted to ~12 mL into 2X storage buffer (80 mM HEPES pH 7.5, 200 mM KCl, 16 mM DTT per ref. (53); 1X listed in **Supp. Table S5E**), and concentrated again to ~0.5 mL. The protein was diluted 1:1 with sterile glycerol (**Supp. Table S5E**), gently mixed, distributed into thirty-eight 20-μL aliquots, and stored at −20°C. The final concentration was evaluated using a Bradford assay against a BSA standard curve. Each aliquot contained ~1 mg/mL of GT-rRI.

Inhibition of RNase A by GT-rRI was assessed by adapting a published method (54), namely, monitoring a reduction in RNase A (Thermo Fisher Scientific# EN0531)-catalyzed hydrolysis of 1 mM cytidine 2’:3’-cyclic monophosphate (cCMP) (Sigma # C9630) in 100 mM Tris-acetate, pH 6.5, 1 mM EDTA, 5 mM DTT at room temperature. Aliquots (18 uL) of a mix containing buffer, DTT, and cCMP were added to Greiner 384-well UV-Star plate (GBO# 788876) wells. RNaseOUT (Thermo Fisher Scientific #10777019), GT-rRI, or water as a positive control for cCMP hydrolysis, was added and allowed to equilibrate in a Synergy™ H4 Hybrid Multi-Mode Microplate Reader for 2 min at 20-25°C before RNase A, 20-100 ng diluted in molecular biology-grade water, or water as negative control for cCMP hydrolysis, was added. Hydrolysis was monitored as the change in absorbance at 286 nm over 30 min.

### Residual RNase activity in purified enzymes

Contaminating RNase activity in purified GT-MMLV and GT*-Taq* was assayed by monitoring degradation of an RNA gel ladder. The reaction solution included the test enzyme at its storage concentration (1 μL), 0.5 μL of Century-Plus RNA Marker (Thermo Fisher Scientific #AM7145), 2 μL of 10X ThermoPol Buffer (NEB #B9004), and nuclease-free water to a final volume of 20 μL. As a positive control, 1 μL of 1 pg/mL RNase A (Thermo Fisher Scientific EN0531) stock solution was added to One*Taq*®. The solutions were incubated at 37°C for 30 min, at which point the reaction was quenched by the addition of an equal volume of 2X loading buffer and dye (95% formamide, 0.025% bromophenol blue, 0.025% xylene cyanol, 5 mM EDTA pH 8.0). The quenched samples were evaluated for ladder degradation by gel electrophoresis using 16 cm x 16 cm 10% polyacrylamide gels with 8 M urea. Gels were run in 1X TBE (Tris/boric acid/EDTA pH 8.0) at 14 W and 300-400 V for at least 30 min prior to loading samples and running for an additional 1.5 h. Gels were stained with ethidium bromide and scanned on the Typhoon Trio+ laser scanner (GE Healthcare) using a channel with 532 nm excitation and a 610 nm emission filter. Smearing of the ladder indicated RNase contamination. The presence of intact bands confirmed the absence of RNase in in-house produced enzymes.

### Formulation and RT-qPCR

1-step RT-qPCR solutions for GT-Master Mix evaluation were prepared in MicroAmp Optical 96-well reaction plates (Applied BioSystems #4346907) and sealed with MicroAmp Optical Adhesive Film (Applied Biosystems, Thermo Fisher Scientific #4311971). Reactions were run on an Applied BioSystems StepOne Plus Real-Time PCR instrument. Except for experiments that tested master mix stability over time, reaction mixtures were prepared by mixing stock reagents immediately prior to conducting the RT-qPCR. The template was Quantitative Synthetic SARS-CoV-2 RNA: ORF, E, N (ATCC #VR-3276S). ROX^TM^ reference dye (Thermo Fisher Scientific #2223012) was included in all reactions for fluorescence normalization. The water used in formulations buffers and reactions was HyClone™ HyPure, Molecular Biology Grade (GE Healthcare #SH30538). Tris-HCl buffers tested ranged from 20-50 mM and pH 8.0-8.8. Monovalent salts (KCl, NaCl, NH_4_ (SO4)_2_) ranged from 10-200 mM. Divalent salts (MgCl_2_, MgSO4) ranged from 1.5-5 mM. Standard dNTPs were included at 0.4-0.5 mM (each). Detergents (0.1% CHAPSO, 0.1% Triton X-100), and other additives (0-1 M betaine, 0-10% trehalose, 0-1 mg/mL bovine serum albumin (BSA, Sigma cat no. A7030)) were tested. Prior to the availability of GT-RT, GT-His-*Taq*, and GT-rRI, commercial materials, namely SuperScript III Reverse Transcriptase (Thermo Fisher Scientific #18080093), Platinum II *Taq* Hot Start DNA Polymerase (Thermo Fisher Scientific #14966001) or *Taq* DNA Polymerase (NEB #M0273), and RNaseOUT (Thermo Fisher Scientific #10777019) or SUPERase•In (Thermo Fisher Scientific #AM2694), were used. IDT N1 primer/probe mix (IDT #10006606), GT multiplex primers/probes, and GT singleplex primers/probes were evaluated. RT-qPCR thermal cycling conditions were modified from those of the CDC protocol (55) (**Table 3**). Unless otherwise noted, the amplification threshold was set to 0.1 and the baseline cycle range was manually set as 3-15 for each run. Once optimal buffer components for GT-produced proteins were identified (**Table 4**), a 2X master mix containing all reaction components except template and primer/probe mix was prepared, tested by RT-qPCR, stored at −20°C, then re-tested by RT-qPCR after sequential freeze-thaw cycles to assess stability and performance over time. PCR efficiency was assessed with Quantitative Synthetic SARS-CoV-2 RNA (ATCC #VR-3276S) or a mixture of full-length SARS-CoV-2 RNA (gift from Dr. Robert Jeffrey Hogan, UGA) and total RNA extracted with TRIzol (Invitrogen #15596026) from HEK293T cells grown to 60% confluency.

### Environmental testing

Environmental surveying was conducted using cotton swabs (Q-tips) as shown in **Figure 4A**. Q-tips were determined experimentally to efficiently collect RNA and viral components from surfaces and not cause RNA degradation. For sample collection one end of the swab was cut off and discarded (**step 1**). The other end of the swab was moistened with 100 μL of “swab medium” (0.5% Triton X-100, 0.05 mM EDTA; **step 2**). The wet swab was placed in a 15 mL Falcon tube and transported to the site to be surveyed. Surface swabbing was conducted in a 6 in^2^ area as shown in **step 3**. After, the swab was returned to the Falcon tube which was tightly closed (**step 4**) and transported on ice to the laboratory. Samples were either processed immediately or stored at −20°C. For processing, the 15-mL Falcon tube containing the swab was incubated at 95°C on a heat block for 5 min to inactivate any live virus that was present (**step 5**). An additional 200-300 μL of swab solution was added to the tube (**step 6**). The tube was vortexed for 10 s (**step 7**), and centrifuged (1000 × g, 2 min, 4°C; **step 8**) prior to analysis by RT-qPCR.

Each 20 μL RT-qPCR survey reaction was assembled using TaqPath 1-Step RT-qPCR Master Mix (Thermo Fisher Scientific #A15300) and 2019-nCoV CDC EUA Kit (primers and probes, IDT #10006606) and analyzed on a StepOne Plus Real-Time PCR instrument (Applied BioSystems) as described above. Samples from swabs were assayed by adding 5 μL of the post survey swab medium directly to the 15 μL RT-qPCR mix. Absolute quantification to determine contaminant copy number in each environmental sample was achieved by comparison to standard curves generated from viral RNA (Quantitative Synthetic SARS-CoV-2 RNA; ATCC #VR-3276SD) or plasmid DNA (2019-nCoV_N_Positive Control; IDT #10006625) (**Fig. 4B**). The amplification threshold was set to 0.1 and the baseline cycle range was manually set as 3-15 for each run. To differentiate between DNA and RNA in samples, the master mix was preheated to 95°C for 5 min (to inactivate the reverse transcriptase) prior to environmental sample addition and cycling.

### Data availability

Raw data available upon request. Plasmids are available except for from us or the original source.

## Data Availability

Protocols available on request

## Acknowledgments

We are grateful for helpful conversations with Andrew Ellington, Amy Lee, Sanchita Bhadra, Greg Gibson, Andre Maranhao, Phil Santangelo, M.G. Finn, Zoe Pratte, Dustin Huard, Moran Frenkel-Pinter, Shweta Biliya, Naima Djeddar, Catherine Moore, Robert Lanciotti, and covidtestingscaleup.slack.com. We acknowledge the core facilities at the Parker H. Petit Institute for Bioengineering and Bioscience at the Georgia Institute of Technology for the use of their shared equipment, services and expertise.

## Funding and additional information

Major funding was provided by the State of Georgia COVID19 Testing Task Force Method Development and Supply Chain Stabilization Studies Proposal (COVID-19 Tech Support Group) and Georgia Institute of Technology. This work was also supported by NASA grants 80NSSC18K1139 and 80NSSC19K0477. RGM and WAM were supported by NIH U54EB027690. The content is solely the responsibility of the authors and does not necessarily represent the official views of the National Institutes of Health.

### Conflict of interest

The authors declare that they have no conflicts of interest with the contents of this article.

Supporting References (56-58)

## References

1. Wu, F., Zhao, S., Yu, B., Chen, Y.-M., Wang, W., Song, Z.-G., Hu, Y., Tao, Z.-W., Tian, J.-H., and Pei, Y.-Y. (2020) A new coronavirus associated with human respiratory disease in China. Nature 579, 265–269

2. Zhou, P., Yang, X.-L., Wang, X.-G., Hu, B., Zhang, L., Zhang, W., Si, H.-R., Zhu, Y., Li, B., and Huang, C.-L. (2020) A pneumonia outbreak associated with a new coronavirus of probable bat origin. Nature 579, 270–273

3. Zhu, N., Zhang, D., Wang, W., Li, X., Yang, B., Song, J., Zhao, X., Huang, B., Shi, W., and Lu, R. (2020) A novel coronavirus from patients with pneumonia in China, 2019. New Engl J Med 382, 727–733

4. Patel, R., Babady, E., Theel, E. S., Storch, G. A., Pinsky, B. A., George, K. S., Smith, T. C., and Bertuzzi, S. (2020) Report from the American Society for Microbiology COVID-19 international summit, 23 march 2020: value of diagnostic testing for SARS–CoV-2/COVID-19. mBio 11, e00722–00720

5. Bhadra, S., Maranhao, A. C., and Ellington, A. D. (2020) One enzyme reverse transcription qPCR using Taq DNA polymerase. *bioRxiv* https://doi.org/10.1101/2020.05.27.120238

6. Graham, T. G. W., Dailey, G. M., Dugast-Darzacq, C., and Esbin, M. N. (2020) BEARmix Version 2. Basic Economical Amplification Reaction one-step RT-qPCR master mix. https://gitlab.com/tjian-darzacq-lab/bearmix/-/raw/master/BEARmix_v2.pdf?inline=false

7. Amen, A. M., Barry, K. W., Boyle, J. M., and Testing Consortium, I. (2020) Blueprint for a pop-up SARS-CoV-2 testing lab. Nat. Biotech. 38, 791–797

8. Holland, P. M., Abramson, R. D., Watson, R., and Gelfand, D. H. (1991) Detection of specific polymerase chain reaction product by utilizing the 5’----3’exonuclease activity of Thermus aquaticus DNA polymerase. Proc Natl Acad Sci USA 88, 7276–7280

9. Esbin, M. N., Whitney, O. N., Chong, S., Maurer, A., Darzacq, X., and Tjian, R. (2020) Overcoming the bottleneck to widespread testing: A rapid review of nucleic acid testing approaches for COVID-19 detection. RNA 26, 771–783

10. Lu, X., Wang, L., Sakthivel, S. K., Whitaker, B., Murray, J., Kamili, S., Lynch, B., Malapati, L., Burke, S. A., and Harcourt, J. (2020) US CDC Real-Time Reverse Transcription PCR Panel for Detection of Severe Acute Respiratory Syndrome Coronavirus 2. Emerging Infect Dis 26, 1654–1665

11. Willman, D. (2020) Contamination at CDC lab delayed rollout of coronavirus tests. *The Washington Post* May 18, 2020, https://www.washingtonpost.com/investigations/contamination-at-cdc-lab-delayed-rollout-of-coronavirus-tests/2020/2004/2018/fd2027d3824-7139-2011ea-aa2080-c2470c2026b2034_story.html

12. Green, M. R., and Sambrook, J. (2018) Hot start polymerase chain reaction (PCR). Cold Spring Harbor Protoc 2018, p.pdb.prot095125

13. (2020) OPTI SARS-CoV-2 RT-PCR Test. https://www.fda.gov/media/137739/download

14. Yeung, A. T., Holloway, B. P., Adams, P. S., and Shipley, G. L. (2004) Evaluation of dual-labeled fluorescent DNA probe purity versus performance in real-time PCR. BioTechniques 36, 266–275

15. Arezi, B., and Hogrefe, H. (2009) Novel mutations in Moloney Murine Leukemia Virus reverse transcriptase increase thermostability through tighter binding to template-primer. Nucleic Acids Res 37, 473–481

16. Bhadra, S., Maranhao, A. C., and Ellington, A. D. (2020) A one-enzyme RT-qPCR assay for SARS-CoV-2, and procedures for reagent production. *bioRxiv* https://doi.org/10.1101/2020.03.29/013342

17. Ellefson, J. W., Gollihar, J., Shroff, R., Shivram, H., Iyer, V. R., and Ellington, A. D. (2016) Synthetic evolutionary origin of a proofreading reverse transcriptase. Science 352, 1590–1593

18. Das, D., and Georgiadis, M. M. (2004) The crystal structure of the monomeric reverse transcriptase from Moloney murine leukemia virus. Structure 12, 819–829

19. Desai, U. J., and Pfaffle, P. K. (1995) Single-step purification of a thermostable DNA polymerase expressed in *Escherichia coli*. BioTechniques 19, 780–782

20. Engelke, D. R., Krikos, A., Bruck, M. E., and Ginsburg, D. (1990) Purification of Thermus aquaticus DNA polymerase expressed in *Escherichia coli*. Anal Biochem 191, 396–400

21. Wang, Y., Prosen, D. E., Mei, L., Sullivan, J. C., Finney, M., and Vander Horn, P. B. (2004) A novel strategy to engineer DNA polymerases for enhanced processivity and improved performance in vitro. Nucleic Acids Res 32, 1197–1207

22. Roux, K. H. (2009) Optimization and troubleshooting in PCR. Cold Spring Harb Protoc 2009, p.pdb.ip66-pdb.ip66

23. Chou, Q., Russell, M., Birch, D. E., Raymond, J., and Bloch, W. (1992) Prevention of pre-PCR mis-priming and primer dimerization improves low-copy-number amplifications. Nucleic Acids Res 20, 1717–1723

24. Davalieva, K., and Efremov, D. G. (2009) Substitution of Ile(707) for Leu in Klentaq DNA polymerase reduces the amplification capacity of the enzyme. Prilozi 30, 57–69

25. Hofsteenge, J., Kieffer, B., Matthies, R., Hemmings, B. A., and Stone, S. R. (1988) Amino acid sequence of the ribonuclease inhibitor from porcine liver reveals the presence of leucine-rich repeats. Biochemistry 27, 8537–8544

26. Kobe, B., and Deisenhofer, J. (1995) A structural basis of the interactions between leucine-rich repeats and protein ligands. Nature 374, 183–186

27. Klink, T. A., Vicentini, A. M., Hofsteenge, J., and Raines, R. T. (2001) High-level soluble production and characterization of porcine ribonuclease inhibitor. Protein Expr Purif 22, 174–179

28. Siurkus, J., and Neubauer, P. (2011) Heterologous production of active ribonuclease inhibitor in *Escherichia coli* by redox state control and chaperonin coexpression. Microb Cell Fact 10, 65

29. Siurkus, J., and Neubauer, P. (2011) Reducing conditions are the key for efficient production of active ribonuclease inhibitor in *Escherichia coli*. Microb Cell Fact 10, 31

30. Sellner, L. N., Coelen, R. J., and Mackenzie, J. S. (1992) Reverse transcriptase inhibits Taq polymerase activity. Nucleic Acids Res 20, 1487–1490

31. Bachmann, B., Luke, W., and Hunsmann, G. (1990) Improvement of PCR amplified DNA sequencing with the aid of detergents. Nucleic Acids Res 18, 1309

32. Farell, E. M., and Alexandre, G. (2012) Bovine serum albumin further enhances the effects of organic solvents on increased yield of polymerase chain reaction of GC-rich templates. BMC Res Notes 5, 257

33. Nagai, M., Yoshida, A., and Sato, N. (1998) Additive effects of bovine serum albumin, dithiothreitol, and glycerol on PCR. Biochem Mol Biol Int 44, 157–163

34. Arakawa, T., and Timasheff, S. N. (1985) The stabilization of proteins by osmolytes. Biophys J 47, 411–414

35. Jain, N. K., and Roy, I. (2009) Effect of trehalose on protein structure. Protein Sci 18, 24–36

36. Beltrán-Pavez, C., Márquez, C. L., Muñoz, G., Valiente-Echeverría, F., Gaggero, A., Soto-Rifo, R., and Barriga, G. P. (2020) SARS-CoV-2 detection from nasopharyngeal swab samples without RNA extraction. *bioRxiv* doi.org/10.1101/2020.03.28.013508.

37. Smyrlaki, I., Ekman, M., Vondracek, M., Papanicoloau, N., Lentini, A., Aarum, J., Muradrasoli, S., Albert, J., Högberg, B., and Reinius, B. (2020) Massive and rapid COVID-19 testing is feasible by extraction-free SARS-CoV-2 RT-qPCR. *medRxiv* https://doi.org/10.1101/2020.04.17.20067348

38. Arumugam, A., and Wong, S. S. (2020) The Potential Use of Unprocessed Sample for RT-qPCR Detection of COVID-19 without an RNA Extraction Step. *bioRxiv*

39. Moore, C., Corden, S., Sinha, J., and Jones, R. (2008) Dry cotton or flocked respiratory swabs as a simple collection technique for the molecular detection of respiratory viruses using real-time NASBA. J Virol Methods 153, 84–89

40. Park, G. W., Chhabra, P., and Vinjé, J. (2017) Swab sampling method for the detection of human norovirus on surfaces. *JoVE*, e55205

41. Kuusela, S., and Lönnberg, H. (1998) Catalytically significant macrochelate formation in Zn2+ promoted hydrolysis of oligoribonucleotides: model studies with chimeric phosphodiester/methylphosphonate oligomers. Nucleos Nucleot Nucl 17, 2417–2427

42. Huff, J. W., Sastry, K. S., Gordon, M. P., and Wacker, W. E. (1964) The action of metal ions on tobacco mosaic virus ribonucleic acid. Biochemistry 3, 501–506

43. Lopez, G. (2020) Why America’s coronavirus testing barely improved in April. *Vox* May 1, 2020, https://www.vox.com/2020/2025/2021/21242589/coronavirus-testing-swab-reagent-supply-shortage

44. Madrigal, A. C., and Meyer, R. (2020) A dire warning from COVID-19 test providers. *Atlantic* June 30, 2020, https://www.theatlantic.com/science/archive/2020/2006/us-coronavirus-testing-could-fail-again/613675/

45. Taylor, S., Wakem, M., Dijkman, G., Alsarraj, M., and Nguyen, M. (2010) A practical approach to RT-qPCR—publishing data that conform to the MIQE guidelines. Methods 50, S1–S5

46. Jones, E., Cooper, A., Bloom, J., Lubock, N., Simpkins, S., Gasperini, M., and Kosuri, S. (2020) Octant SwabSeq Testing. https://www.notion.so/Octant-SwabSeq-Testing-9eb80e793d797e46348038aa46348080a46348035a46348901fd#46348035cf46347548d46348573a46348034c46348648fc46348092f46348038bb47853208

47. The Francis Crick Institute. https://www.crick.ac.uk/research/covid-19/covid19-consortium

48. de Oliveira Mann, C. C., Orzalli, M. H., King, D. S., Kagan, J. C., Lee, A. S. Y., and Kranzusch, P. J. (2019) Modular Architecture of the STING C-Terminal Tail Allows Interferon and NF-kappaB Signaling Adaptation. Cell Rep 27, 1165-1175 e1165

49. Thapa, H. R., Lin, Z., Yi, D., Smith, J. E., Schmidt, E. W., and Agarwal, V. (2020) Genetic and Biochemical Reconstitution of Bromoform Biosynthesis in Asparagopsis Lends Insights into Seaweed Reactive Oxygen Species Enzymology. ACS Chem Biol 15, 1662–1670

50. Studier, F. W. (2005) Protein production by auto-induction in high density shaking cultures. Protein Expr Purif 41, 207–234

51. Zeng, F., Zhang, S., Hao, Z., Duan, S., Meng, Y., Li, P., Dong, J., and Lin, Y. (2018) Efficient strategy for introducing large and multiple changes in plasmid DNA. Sci Rep 8, 1714–1726

52. Hill, S. E., Donegan, R. K., Nguyen, E., Desai, T. M., and Lieberman, R. L. (2015) Molecular Details of Olfactomedin Domains Provide Pathway to Structure-Function Studies. PLoS One 10, e0130888

53. Guo, W., Cao, L., Jia, Z., Wu, G., Li, T., Lu, F., and Lu, Z. (2011) High level soluble production of functional ribonuclease inhibitor in *Escherichia coli* by fusing it to soluble partners. Protein Expr Purif 77, 185–192

54. Blackburn, P. (1979) Ribonuclease inhibitor from human placenta: rapid purification and assay. J Biol Chem 254, 12484–12487

55. CDC. (March 15, 2020) 2019-Novel Coronavirus (2019-nCoV) Real-Time RT-PCR Diagnostic Panel, Instructions for use. CDC-006–00019, Revision: 02.

56. Dang, C., and Jayasena, S. D. (1996) Oligonucleotide inhibitors of TaqDNA polymerase facilitate detection of low copy number targets by PCR. J Mol Biol 264, 268–278

57. Gold, L., and Jayasena, S. D. (1997) Nucleic acid ligand inhibitors to DNA polymerases. in *US6183967B1*, Google Patents

58. Lin, Y., and Jayasena, S. D. (1997) Inhibition of multiple thermostable DNA polymerases by a heterodimeric aptamer. J Mol Biol 271, 100–111

